# Depicting the regulatory role of JZOL on TRP channels in the treatment of Acute Bronchitis based on the combination of clinical trials, computational analysis and in vivo experiments

**DOI:** 10.1101/2024.05.07.24306993

**Authors:** Qinhua Fan, Chongming Wu, Yawei Du, Boyang Wang, Yanming Xie, Zeling Zhang, Wenquan Su, Zizhuo Wang, Changchang Xu, Xueke Li, Ying Ding, Xinjiang An, Jing Chen, Yunying Xiao, Rong Yu, Nan Li, Juan Wang, Yiqun Teng, Hongfen Lv, Nian Yang, Yuling Wen, Xiaoli Huang, Wei Pan, Yufeng Liu, Xueqin Xi, Qianye Zhao, Changshan Liu, Jian Xu, Haitao Zhang, Lie Zhuo, Qiangquan Rong, Yu Xia, Qin Shen, Shao Li, Junhong Wang, Shengxian Wu

## Abstract

The comparison between traditional Chinese medicine Jinzhen Oral Liquid (JZOL) and western medicine in treating children with acute bronchitis (AB) showed encouraging outcomes. This trial evaluated the efficacy and safety of the JZOL for improving cough and expectoration in children with AB. 480 children were randomly assigned to take JZOL or Ambroxol Hydrochloride and Clenbuterol Hydrochloride Oral Solution for 7 days. The primary outcome was time-to-cough resolution. The median time-to-cough resolution in both groups was 5.0 days and the antitussive onset median time was only 1 day. This head to head randomized controlled trial showed that JZOL was not inferior to cough suppressant and phlegm resolving western medicine in treating cough and sputum and could comprehensively treat respiratory and systemic discomfort symptoms. Combined with clinical trials, the mechanism of JZOL against AB was uncovered by network target analysis, it was found that the pathways in TRP channels like IL-1β/IL1R/TRPV1/TRPA1, NGF/TrkA/TRPV1/TRPA1 and PGE2/EP/PKA/TRPV1/TRPA1 might play important roles. Animal experiments further confirmed that inflammation and immune regulatory effect of JZOL in the treatment of AB were of vital importance and TRP channels was the key mechanism of action.

## 1. Introduction

Acute bronchitis (AB) is a prevalent respiratory infection among children^1^, and is a leading cause of hospitalization for children^2^. Approximately one-third of children visit pediatricians due to AB^3^, accounting for 40% of all healthcare visits in child care institutions^4^. Currently, there is no specific therapeutic medication for AB except symptomatic supportive treatment. Antibiotics are commonly prescribed as the initial treatment in western medicine^5^, but the applicability and effectiveness have been widely disputed. Cough relieving and expectorant medicines can inhibit the central nervous system^6^, the US Food and Drug Administration even advised not to use cough preparations for children under six^7^. Although there are many western medicines, the range of pediatric options is limited. The clinical drugs for treating AB is lack and urgent at present.

Complementary and Alternative Medicine (CAM) has been widely used in pediatrics, and parents generally believe that CAM is “natural”^8,9^. It has been reported that more than 50% of Canadian and European children received CAM treatment during illness^10–12^. Traditional Chinese medicine (TCM) has become an important CAM approach, with good therapeutic advantages for respiratory diseases^13^. Based on the clinical characteristics of AB, cough, fever, or dry mouth, TCM categorizes AB as “phlegm heat syndrome”, representing bronchitis in the acute stage in TCM theory. Jinzhen Oral Liquid (JZOL), a TCM compound formulation, is widely used in pediatric clinics in China and is highly recognized for its significant therapeutic effect on children with AB. JZOL prescription was derived from the experience of TCM in preventing and treating acute respiratory infections and is suitable for the “phlegm heat syndrome”^14–16^. Using UPLC-Q/TOF-MS technology analysis, the main components absorbed into the bloodstream included flavonoids, triterpenes, and anthraquinones; the main active ingredients were flavonoids represented by baicalin^17^.

In China, JZOL is recommended for acute respiratory infections in children^18^. *In vitro* and *in vivo* studies have substantiated that JZOL has anti-inflammatory and broad-spectrum antiviral effects and has potential therapeutic effects on AB^19–22^. Some studies about JZOL have been published in China; these early trials all reported that JZOL yielded significant benefits in improving symptoms in AB, but with the low quality of research conducted. However, most clinical trials adopted western medicine combined with JZOL compared with western medicine alone, there was no solid evidence to certificate the beneficial functions of JZOL in treating AB. This study was the first head to head prospective clinical trial of JZOL in treating children with AB strictly according to the CONSORT Extension for CHM Formulas^23^, aimed to evaluate the efficacy and safety of the JZOL.

In the current study, we conducted a prospective clinical trial of JZOL in treating children with AB, which confirmed its therapeutic effect. We further performed network target analysis^24,25^ to predict targets profiles of each compounds in JZOL. Based on the network target analysis, we conducted animal experiments to confirm the therapeutic effect and mechanism of JZOL and further validate its mechanism.

## 2. Methods and Materials

### 2.1 Clinical trial

#### 2.1.1 Design Overview

A non-inferiority, randomized, double-blind, double-simulated, multicenter clinical trial was carried out in 24 hospitals in different cities across the country in China. The study has been approved by the Ethics Committee of Dongzhimen Hospital, affiliated with the Beijing University of Chinese Medicine and each branch center (September 16, 2019, v1.1, No. DZMEC-KY-2019-183). Compared to the published protocol^26^, this study increased the number of branch centers to 24 hospitals, changed the sample size to 480 cases (v1.3, No. DZMEC-KY-2019-183-02), and conducted network target analysis and animal experiments, instead of collecting blood, saliva, and fecal samples, for mechanism research. All participants provided informed consent. This study has been registered with the Chinese Clinical Trial Registry (ChiCTR200034703).

#### 2.1.2 Patient Enrollment

Pediatricians established the diagnosis of AB according to clinical manifestations, physical examination, X-ray, and laboratory examination. We included children with AB aged 2-14 years (≥2 years, <15 years) and with a course of less than 48 hours. Besides, all included participants had cough (day + night) scores ≥4 points and took no antibiotics, antitussives, phlegm-reducing drugs, or other medication that may influence cough before treatment.

We excluded patients who with severe bronchitis, which was difficult to distinguish from pneumonia in the early stage, those with acute infectious diseases such as measles, pertussis, and influenza, patients with single acute upper respiratory tract infections, purulent tonsillitis, asthmatic bronchitis, bronchial asthma, bronchiolitis pneumonia, tuberculosis, or tumor, a large amount of purulent sputum in the lungs, severe malnutrition or immune deficiency, severe primary diseases including heart, liver, kidney, digestive and hematopoietic systems. Patients were also excluded if they were allergic to JZOL or Ambroxol Hydrochloride and Clenbuterol Hydrochloride Oral Solution (AHCHOS), were receiving epinephrine, isoproterenol or other catecholamines, monoamine oxidase inhibitors or tricyclic antidepressants, propranolol or other nonselective β-blockers, large amounts of other sympathetic stimulants.

The informed consent forms were jointly signed by the legal guardian and/or child (≥8 years old).

#### 2.1.3 Randomization and Blinding

This clinical trial adopted the stratified block randomization method. Patients were stratified according to different age groups (2-3 years old, 4-5 years old, 6-7 years old, 8-12 years old, and >12 years old), and each sub-center participated in the selection process. Using SAS 9.4 to generate random numbers, random numbers were assigned through the Clinical Trial Central Randomization System (DAS for IWRS). Participants were assigned randomly to the intervention and control groups in a 1:1 ratio. The drug blinding was carried out by staff members from the statistical unit and applicant unit, who had no connection to this trial. As a result, both the researchers and participants were kept unaware of the drug allocation.

#### 2.1.4 Interventions

JZOL was produced by Jiangsu Kangyuan Pharmaceutical Co., Ltd. **Table 1** lists the specific composition and content. AHCHOS (every 100ml of AHCHOS contains ambroxol hydrochloride 150mg and clenbuterol hydrochloride 0.1mg) was produced by Beijing Hanmei Pharmaceutical Co., Ltd. JZOL placebo, and AHCHOS placebo was provided by Jiangsu Kanion Pharmaceutical Co., Ltd according to the double-blind principle. The emergency antipyretic and analgesic were ibuprofen suspension (Shanghai Johnson & Johnson Pharmaceutical Co., Ltd). All drugs met quality standards.

**Table 1.**
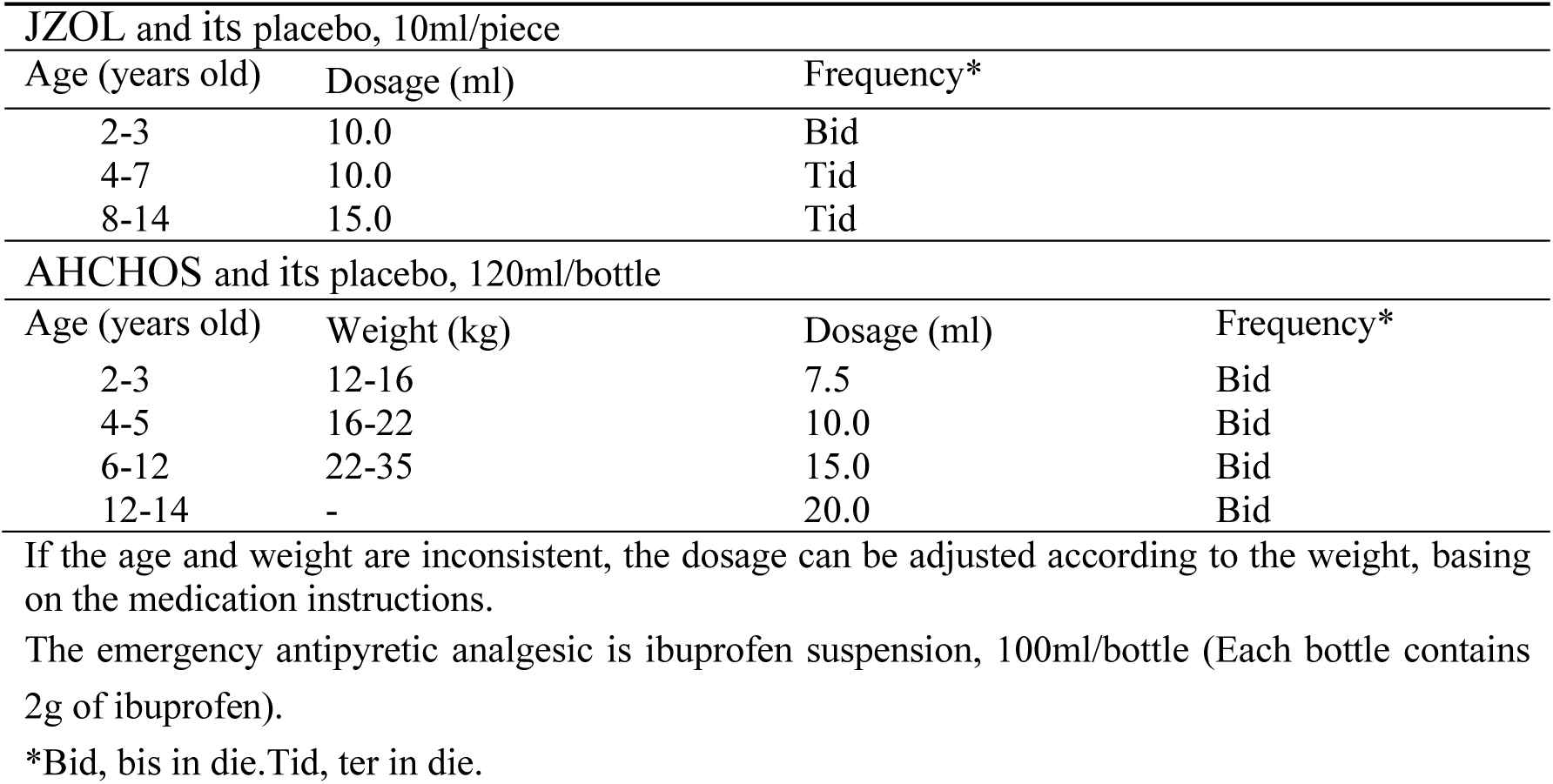
Composition of Jinzhen Oral Liquid.

During the 7-day medication period, individuals in the intervention group were administered JZOL and AHCHOS placebo concurrently. In contrast, those in the control group were given AHCHOS and JZOL placebo. **Table 2** lists the usage instructions.

**Table 2.**
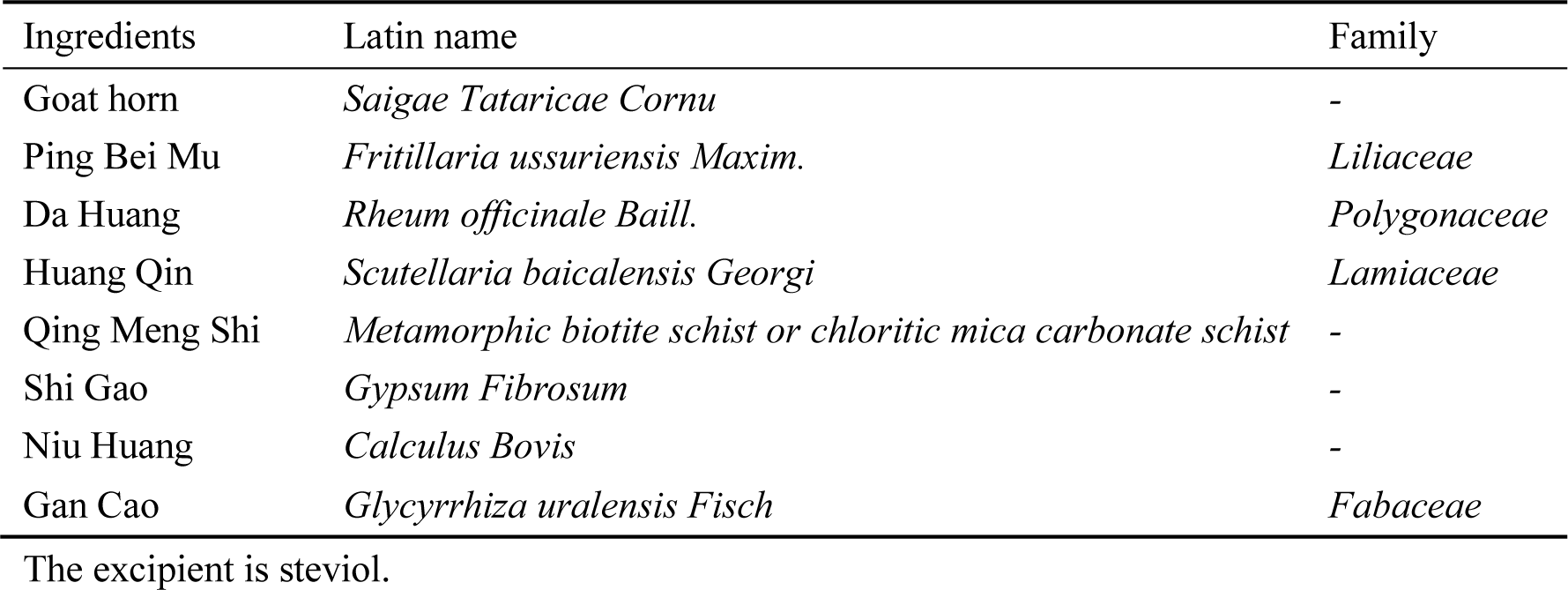
Usage instructions for both drugs.

During the trial, participants were prohibited from using expectorants, bronchodilators, or other medications with cough-relieving and expectorant effects. Routine administration of antibiotics was not provided; however, they could be given if necessary by a medical professional. If participants’ axillary temperature exceeded 38.5℃ during the trial, they could use physical cooling or antipyretic and analgesic medication (such as ibuprofen suspension) under medical guidance. Any combined medications used were meticulously documented.

#### 2.1.5 Assessment

During the clinical trial, patients’ cough scores were recorded daily (0=absent, 1=mill, 2=moderate, and 3=severe). The primary outcome was time-to-cough resolution (day + night cough scores ≤1 point, lasting for 24 hours). The secondary outcomes included cough score (total scores of day and night cough scores), onset time of antitussive action (the number of days required for cough scores to decrease by 1 point after treatment), Area Under Curve (AUC) of cough and expectoration symptom score-time, clinical recovery time (the number of days required for recovery after treatment when cough, expectoration, and lung signs were mild and didn’t affect learning, life, and sleep). The above indicators were recorded daily and evaluated at the end of the study. Besides, we recorded the disappearance rate of secondary clinical symptoms (fever, thirst, red-faced, dysphoria, short voidings of reddish urine, and dry stool) about AB, normalization situation of abnormal white blood cell count, lymphocyte percentage, and neutrophil percentage^27–30^, the disappearance rate of cough, sputum and pulmonary rales and the usage of antibiotics, antipyretics and analgesics. These indicators were compared with the baseline at the end of the study.

### 2.2 Network target analysis

Targets profiles of each compounds in JZOL were predicted by a network-based algorithm drugCIPHER^31,32^. Further the holistic targets of JZOL were constructed based on our previous statistical model^33,34^. Enrichment of Kyoto Encyclopedia of Genes and Genomes (KEGG) and Gene Ontology (GO) were performed on the holistic targets of JZOL to shift our focus from the target level to the pathways or biological processes with R package clusterProfiler v4.9.1. Modular networks were constructed based on a high-quality protein-protein interactome and visualized by Cytoscape v3.8.1.

In order to find the mechanism of JZOL in the treatment of cough, molecules related to cough were collected from CTD database^35^, Gendoo^36^ and DisGenet^37^. Pathways enriched by both the holistic targets of JZOL and molecules related to cough were focused for further investigation. The signaling pathways in the network target of JZOL against cough were depicted based on KEGG.

### 2.3 Animal experiments

In this study, five animal models were constructed, including two models of inflammation in the trachea, one cough hypersensitivity model, one phenol red excretion model and one citrate acid induced cough model. More details were listed in Supplementary materials.

### 2.4 Statistical Analysis

All the data of the clinical trial was analyzed by the third-party statistical agency (Shanghai Bojia Pharmaceutical Technology Co., Ltd). Statistical analysis was used to estimate sample size. The primary outcome was time-to-cough resolution. The expected median time-to-cough resolution in the control group was 5 days, and recruitment time was 18 months. We observed all patients until day 7 of their enrollment. To determine non-inferiority, the hazard ratio (HR) threshold was set at 0.75, with a one-sided α of 0.025 and a power of 0.8. Based on calculations using the PASS14.0 software, a sample size of 480 cases was estimated, with approximately 20% expected dropout rate. This final sample size included 240 cases each in both the intervention and control groups, with 379 events observed.

This clinical study did not conduct a mid-term analysis. Statistical analysis was based on the principle of intention-to-treat (ITT), which included all randomly assigned participants. Analysis based on actual data was performed using the full analysis set (FAS). Continuous variables were analyzed based on mean, standard deviation, median, and quartile, while frequency and composition ratio were applied to ranking and counting data.

The time-to-cough resolution was analyzed using the Kaplan-Meier method, which provided estimates of the median, 25% and 75% percentiles, and 95% confidence intervals (Brookmeyer Crowley method). An inter-group comparison was conducted using the log-rank test, which adjusted for age stratification factors. The inter-group hazard ratio was calculated using a Cox proportional risk regression model, which adjusted for age stratification factors. The HR was also used to determine if the intervention group was non-inferior to the control group, based on a preset HR threshold of 0.75. The 95% confidence interval for the HR was estimated using the Wald method. In addition, we used a log-rank test to analyze the time-to-cough resolution in different age groups (2-3 years old, 4-5 years old, 6-7 years old, 8-12 years old, and >12 years old).

The onset time of antitussive action and clinical recovery time were analyzed using the Kaplan-Meier method, which provided estimates of the median time, 25%, and 75% percentiles, and 95% confidence intervals (Brookmeyer Crowley method), the log-rank test was used for inter-group comparison after adjusting for age. Cough score, AUC of cough and expectoration symptom score-time, white blood cell count, lymphocyte percentage, and neutrophil percentage were analyzed using a t-test. The disappearance rate of secondary clinical symptoms (fever, thirst, red-faced complexion, dysphoria, short voidings of reddish urine, and dry stool), the disappearance rate of cough, sputum and pulmonary rales, the recovery rate of abnormal laboratory indicators (white blood cell count, lymphocyte percentage and neutrophil percentage), and the usage of antibiotics, antipyretics and analgesics were analyzed by the Chi-square test or Fisher exact probability method.

To determine the time-to-cough resolution, onset time of antitussive action, and clinical recovery time, we adopted the longest observation time of all participants as the deletion time. When analyzing the AUC of cough and expectoration symptom score-time, the scores of participants who withdrew from the study early were substituted with their last available cough and expectoration score before withdrawal, up to the 7th day, which was the analysis endpoint. Missing data from the baseline, other efficacy indicators, and safety evaluations were not imputed.

All statistical tests of the clinical trial were conducted using SAS 9.4 software, bilateral tests, *p* < 0.05 was considered statistically significant. The bilateral 95% confidence interval of intergroup differences was used to represent the non-inferiority test results.

The data of animal experiments were analyzed by Graphpad Prism 8.0 software to analyze the data of each group, with continuous data presented as mean ± standard deviation (±SD). Inter-group comparisons were made using ANOVA multiple comparisons or Student’s t-test, with *p* < 0.05 indicating statistical significance.

### 2.5 Public based validation on the levels of compounds and herbs

In order to validate our findings at the level of compounds or herbs, two public transcriptomics datasets were collected from ITCM and a previous study^38,39^. These two datasets included the transcriptomes of common compounds in TCM on MCF-7 cell lines and the transcriptomes of common herbs on THP-1 cell lines, respectively. Transcriptomics data were processed through R package DESeq2 v1.30.0 and differential statistical tests were performed in student’s t test.

## 3. Results

### 3.1 The clinical efficacy and safety of JZOL against AB

#### 3.1.1 Participant Characteristics

Between January 19, 2021 and April 8, 2022, 517 patients aged 2-14 years were screened, out of which 37 cases were excluded. Eventually, 480 participants who met the criteria were randomly assigned, of which 11 participants in the intervention group withdrew early without receiving any intervention. The final study population consisted of 469 subjects (236 subjects in the intervention group and 233 cases in the control group) (**Figure 1**), exhibiting male predominance (54.4%), with an average age of 5.7 (SD, 2.74) years and an average weight of 24.36 (SD, 10.585) kg. The median time from onset to randomization was 36.0 hours (Q1 to Q3, 25.0 to 47.0 hours). **Table 3** lists the baseline characteristics of the subjects.

**Figure 1.**
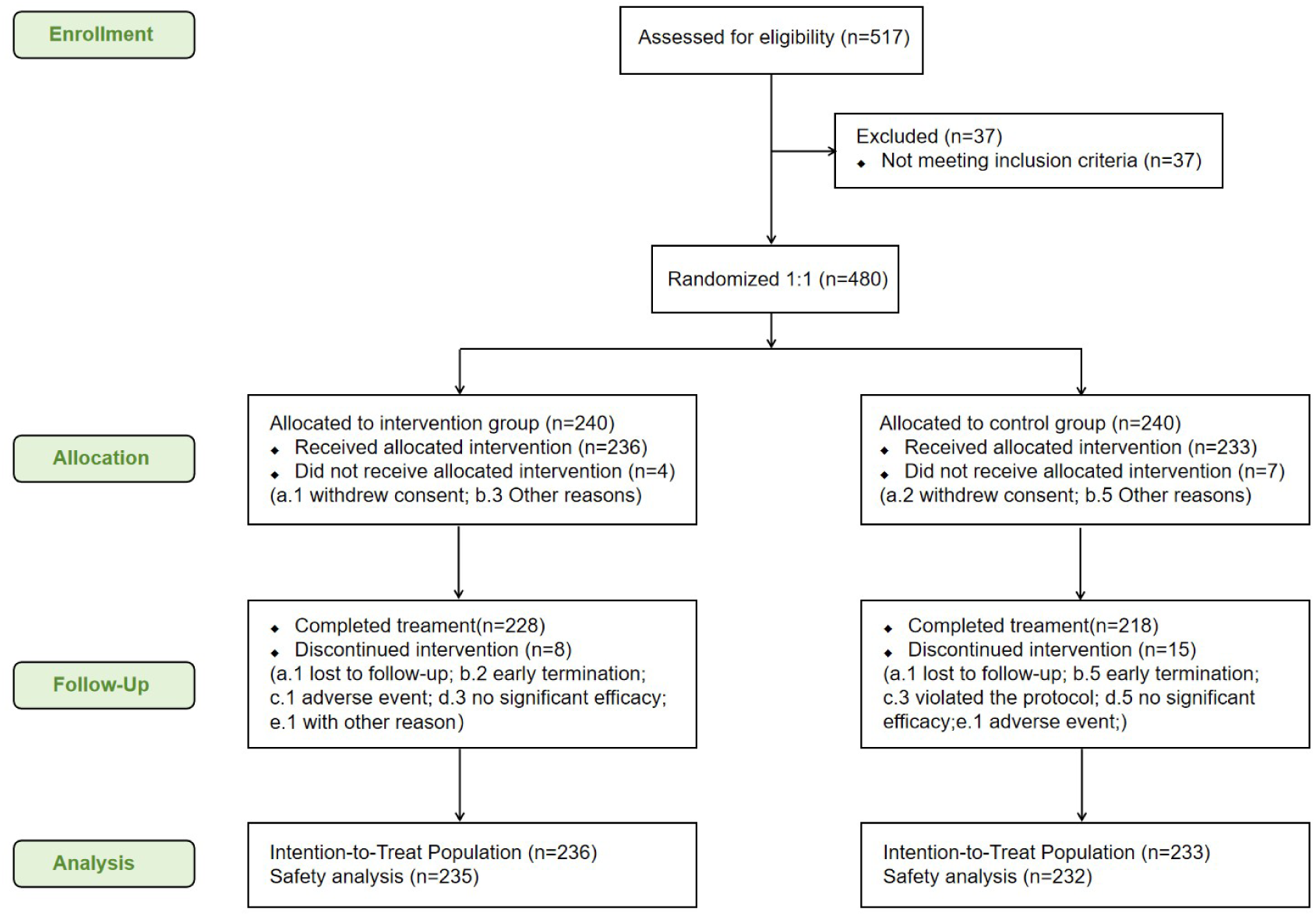
Flow diagram of the clinical trial

**Table 3.**
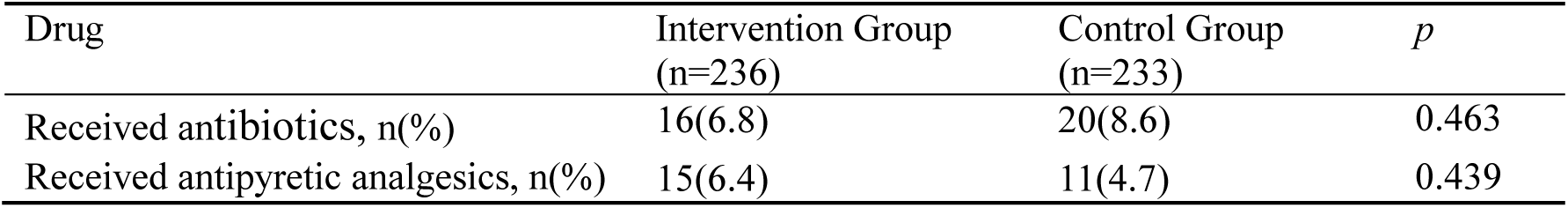
Baseline Characteristics of Patients.

At baseline, neither group was administered antibiotics, although 4 subjects in the intervention group and 3 in the control group took antipyretic analgesic drugs. After enrollment, the usage of antibiotics (6.8% in the intervention group, 8.6% in the control group; *p*=0.463) and antipyretic analgesics (6.4% in the intervention group, 4.8% in the control group; *p*=0.439) in the two groups was similar. The antipyretic analgesic used by patients after admission was ibuprofen suspension, the emergency medication specified in the trial. The antibiotics used in this study mainly included azithromycin, amoxicillin, erythromycin, cephalosporin, etc. **Table 4** shows the usage of antibiotics and antipyretic analgesics after treatment.

**Table 4.**
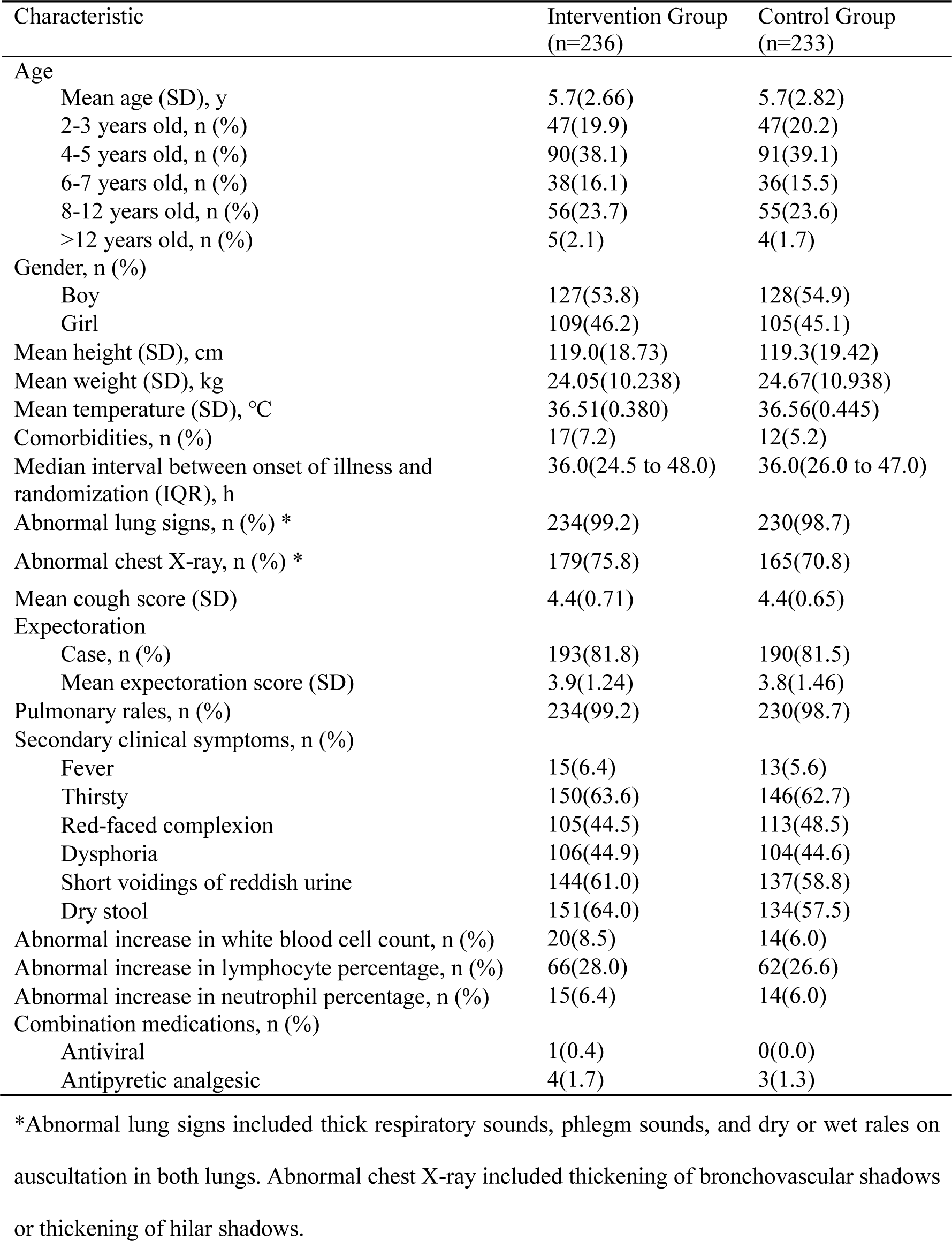
Use of antibiotics and antipyretic analgesics.

#### 3.1.2 Primary clinical outcomes

The median time-to-cough resolution in both the intervention and control groups was 5.0 days (CI, [NA to NA]), and there was no significant difference between the two groups (*p*=0.773). According to the Cox proportional risk regression model, the HR of time-to-cough resolution between the two groups was 0.975 (CI, 0.794 to 1.197), suggesting the intervention group outcomes were not inferior to the control group (**Figure 2**). **Table 5** lists the primary outcome after treatment in two groups.

**Figure 2.**
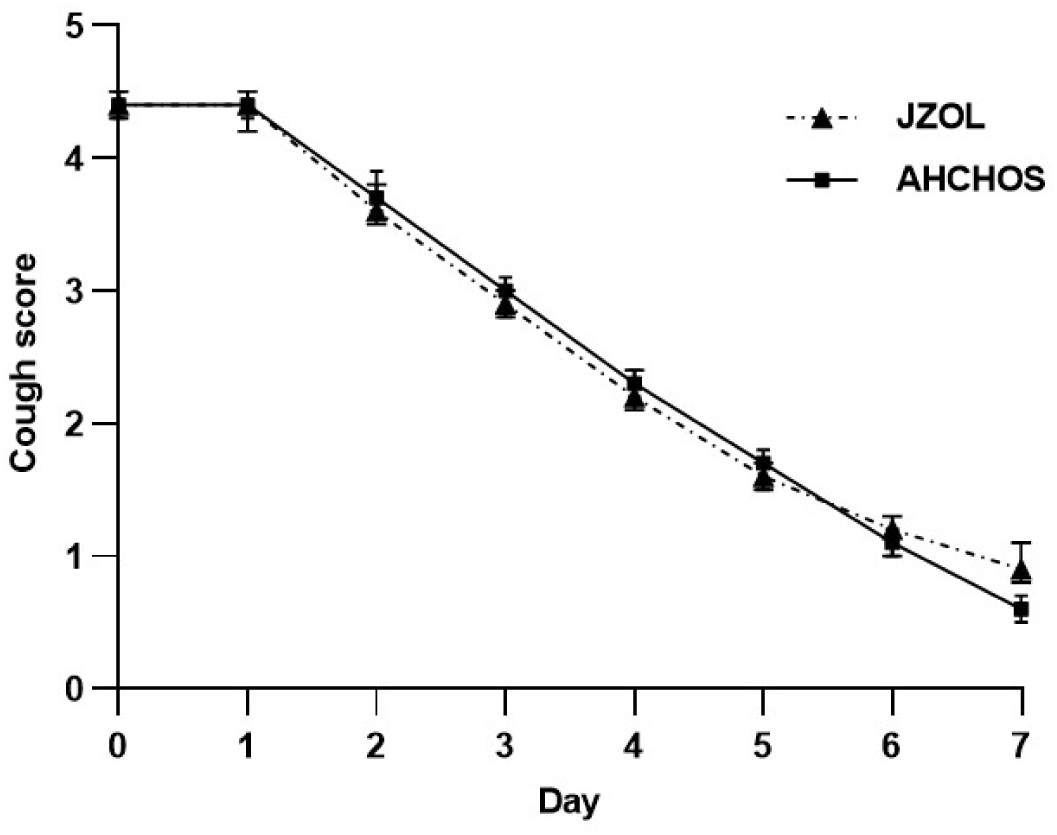
Daily time-to-cough resolution. Error bars represent 95% CIs.

**Table 5.** Outcomes of coughing and clinical recovery time.

| Outcome | Intervention Group<br>(n=236) | Control Group<br>(n=233) | Between-Group<br>Difference (95%<br>CI) | *p* |
| --- | --- | --- | --- | --- |
| Median time-to-cough resolution (95% CI), d* | 5.0(NA to NA) | 5.0(NA to NA) | - | 0.773 |
| median onset time to antitussive action (95% CI), d† | 1.0(1.0 to 2.0) | 1.0(1.0 to 2.0) | - | 0.519 |
| Change in cough score (95% CI) |  |  |  |  |
| Day 1 | -0.1(-0.2 to 0.0) | -0.0(-0.1 to 0.0) | -0.05(-0.13 to 0.02) | 0.153 |
| Day 2 | -0.8(-0.9 to -0.7) | -0.7(-0.8 to -0.5) | -0.17(-0.33 to -0.00) | 0.048 |
| Day 3 | -1.6(-1.7 to -1.4) | -1.4(-1.6 to -1.3) | -0.13(-0.32 to 0.06) | 0.173 |
| Day 4 | -2.2(-2.3 to -2.0) | -2.1(-2.2 to -1.9) | -0.12(-0.33 to 0.08) | 0.248 |
| Day 5 | -2.8(-3.0 to -2.7) | -2.7(-2.9 to -2.6) | -0.09(-0.31 to 0.13) | 0.432 |
| Day 6 | -3.3 (-3.5 to -3.1) | -3.3 (-3.4 to -3.1) | -0.01(-0.23 to 0.22) | 0.958 |
| Day 7 | -3.5 (-3.6 to -3.3) | -3.7 (-3.9 to -3.6) | 0.29(0.03 to 0.55) | 0.029 |
| AUC of cough symptom score-time (95% CI) | 18.66(18.07 to 19.25) | 19.17 (18.50 to 19.84) | -0.51(-1.40 to 0.38) | 0.257 |
| Disappearance rate of cough on Day 7, n (%) ‡ | 180(77.3) | 188(82.5) | -5.2(-12.5 to 2.1) | 0.164 |
| Clinical recovery median time (95% CI), d† | 6.0(5.0 to 6.0) | 6.0(5.0 to 6.0) | - | 0.901 |

\*Calculated *p* using the stratified log-rank test, calculated HR using stratified Cox proportional risk regression model. The stratification factor was age (2-3 years old, 4-5 years old, 6-7 years old, 8-12 years old,>12 years old); the preset non-inferior threshold was 0.75.
†Calculated *p* using stratified log-rank test; the stratification factor was age (2-3 years old, 4-5 years old, 6-7 years old, 8-12 years old,>12 years old).
‡Calculated the disappearance rate of cough after treatment for two groups of patients with cough at baseline. Compared to the baseline, 3 cases were deleted in the intervention group and 5 in the control group.

Different ages were required to take different doses of medication, we conducted a subgroup analysis based on age. **Table 6** lists the time-to-cough resolution in different age groups. The intervention group had the shortest median time for cough disappearance in the age group of 8-12 years, which was 4.5 days (CI, 4.0 d to 5.0 d), while the control group had the shortest median time for cough disappearance in the age group of >12 years, which was 4.0 days (CI, 1.0d to NA). In other age groups, both groups were 5.0 days. There was no significant difference in the median time-to-cough resolution between the two groups at all age groups (in 2-3 years old, 4-5 years old, 6-7 years old, 8-12 years old, >12 years old, *p*=0.091, 0.945, 0.893, 0.267, 0.172).

**Table 6.**
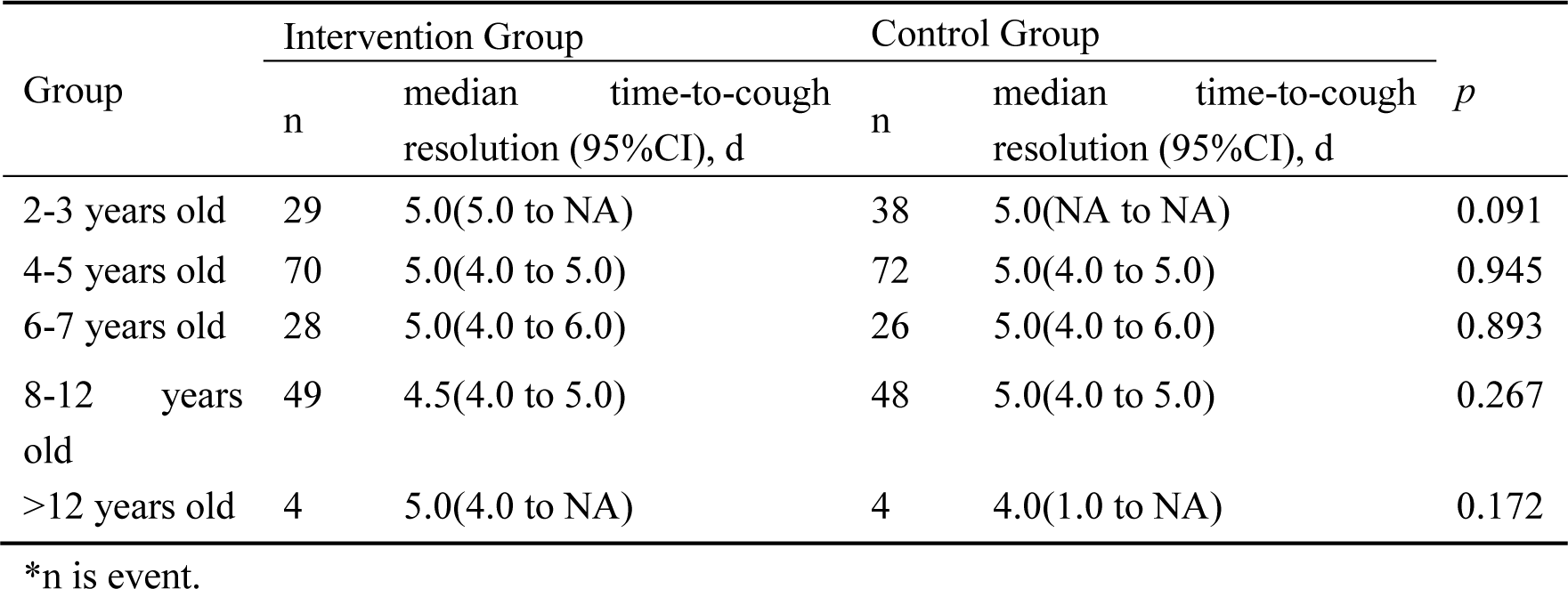
Time-to-cough resolution in different age groups Intervention Group Control Group.

#### 3.1.3 Secondary clinical outcomes

In the average cough score, there was no significant difference between the two groups from day 1 to day 6. On day 7 of treatment, the cough score in the control group decreased by 3.7 (CI, 3.9 to 3.6), which was higher than the decreased score in the intervention group (3.5 [CI, 3.6 to 3.3]) (intergroup difference was 0.29 [CI, 0.03 to 0.55]; *p*=0.029). On day 7, the average cough score (0.6 [CI, 0.5 to 0.7]) in the control group was lower than the intervention group (0.9 [CI, 0.8 to 1.1]) (intergroup difference was 0.31 [CI, 0.13 to 0.50]; *p*<0.001) (**Figures 3**, 4). The median onset time of antitussive action in both the intervention and control groups was 1.0 days (CI, [1.0d to 2.0d]), and the median clinical recovery time was both 6.0 days (CI, [5.0d to 6.0d]). The two groups had no significant differences in median onset time to antitussive action and median clinical recovery time (*p*=0.519 and 0.901, respectively). **Table 5** lists the cough scores after treatment.

**Figure 3.**
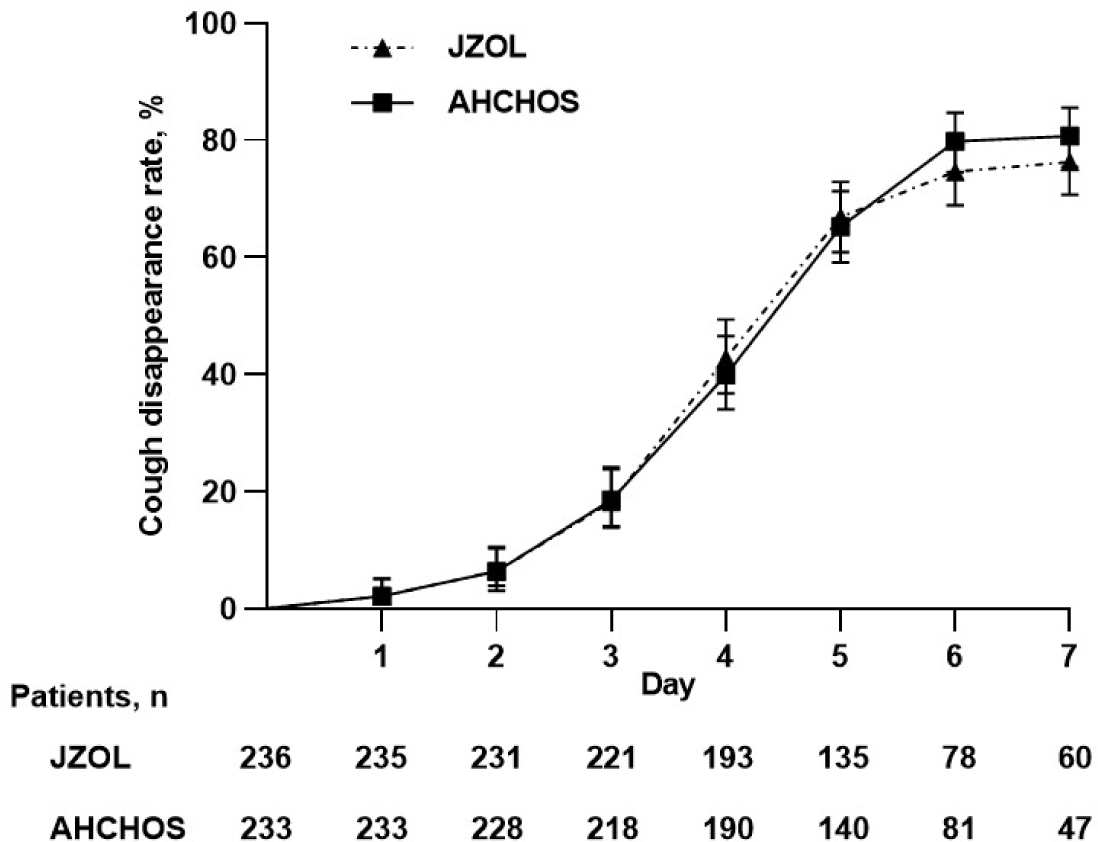
Daily Cough score. Error bars represent 95% CIs.

**Figure 4.**
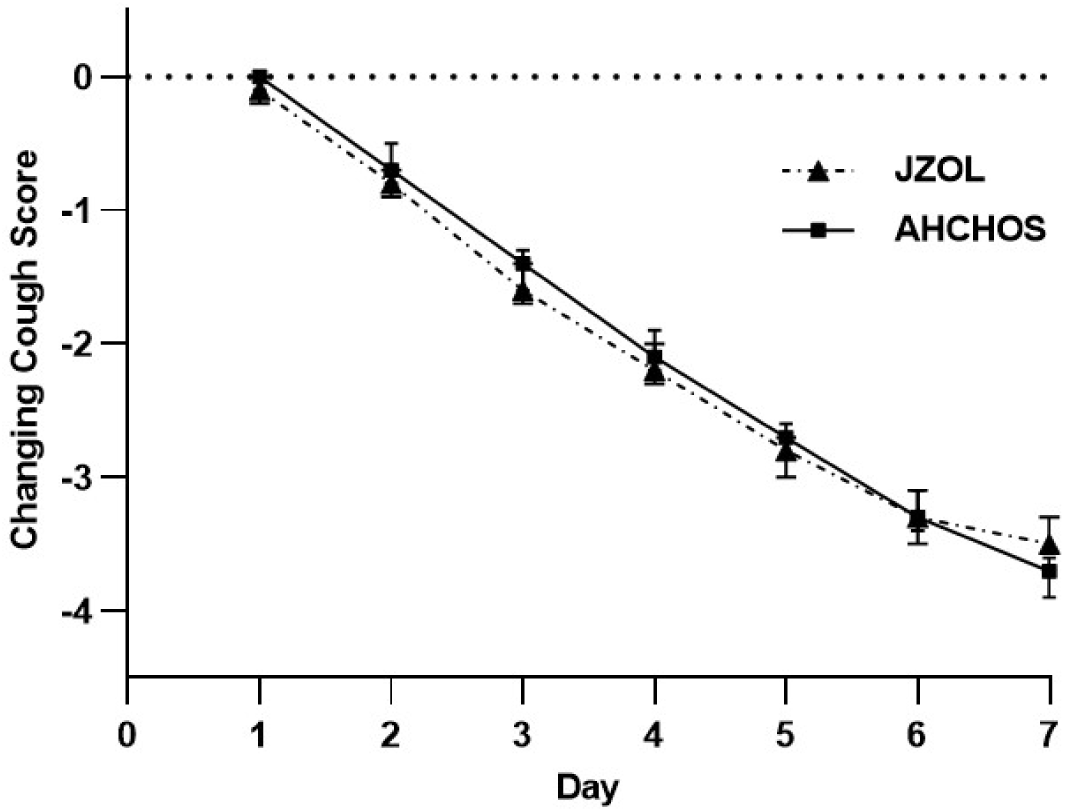
Changes in daily cough score. Error bars represent 95% CIs.

Compared with the control group, the AUC of 0-7d cough and expectoration symptom score-time and the disappearance rate of cough and sputum of the intervention group were similar (*p*=0.257, 0.563, 0.164, 0.905, respectively). However, the disappearance rate of pulmonary rales in the intervention group (96.3%) was significantly higher than in the control group (89.8%) after treatment (*p*=0.037) (**Table 7**).

**Table 7.**
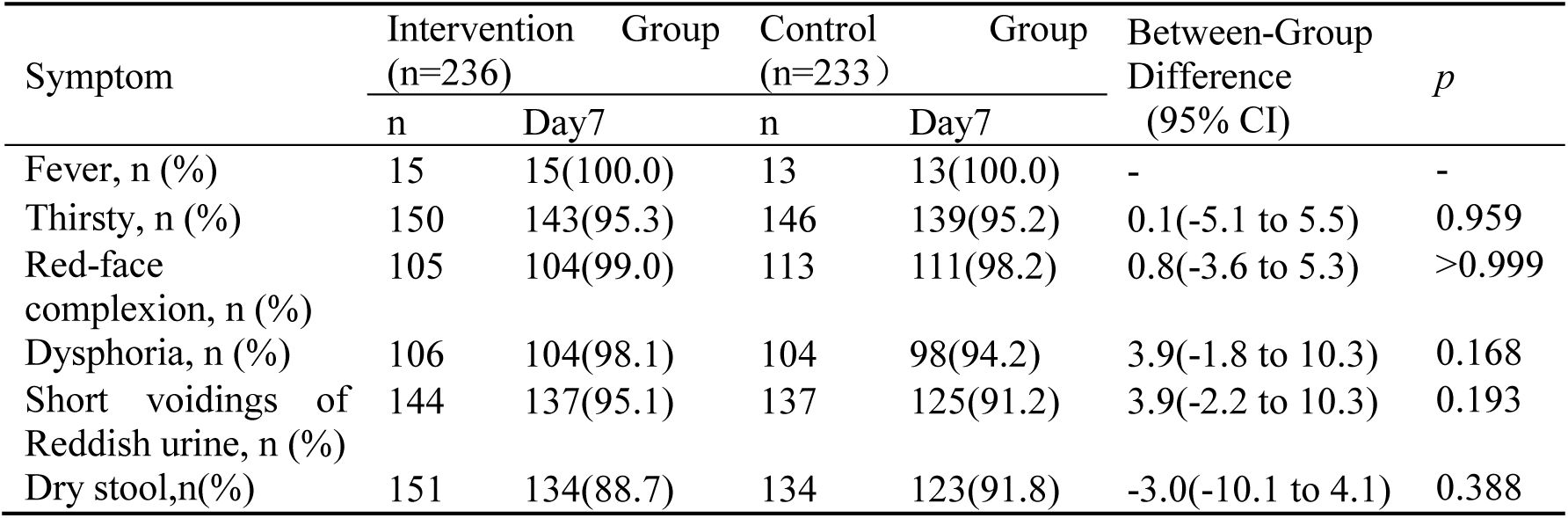
Outcomes of expectoration and pulmonary rales.

In the secondary clinical symptoms in two groups, after treatment, two groups of patients showed significant improvement in fever, thirst, red-faced complexion, dysphoria, short voidings of reddish urine, and dry stool compared to baseline. Although there was no significant difference in the disappearance rate of secondary clinical symptoms between the two groups, the intervention group exhibited slight advantages over the control group related to thirst, red-faced complexion, dysphoria, and short voidings of reddish urine (**Table 8**).

**Table 8.**
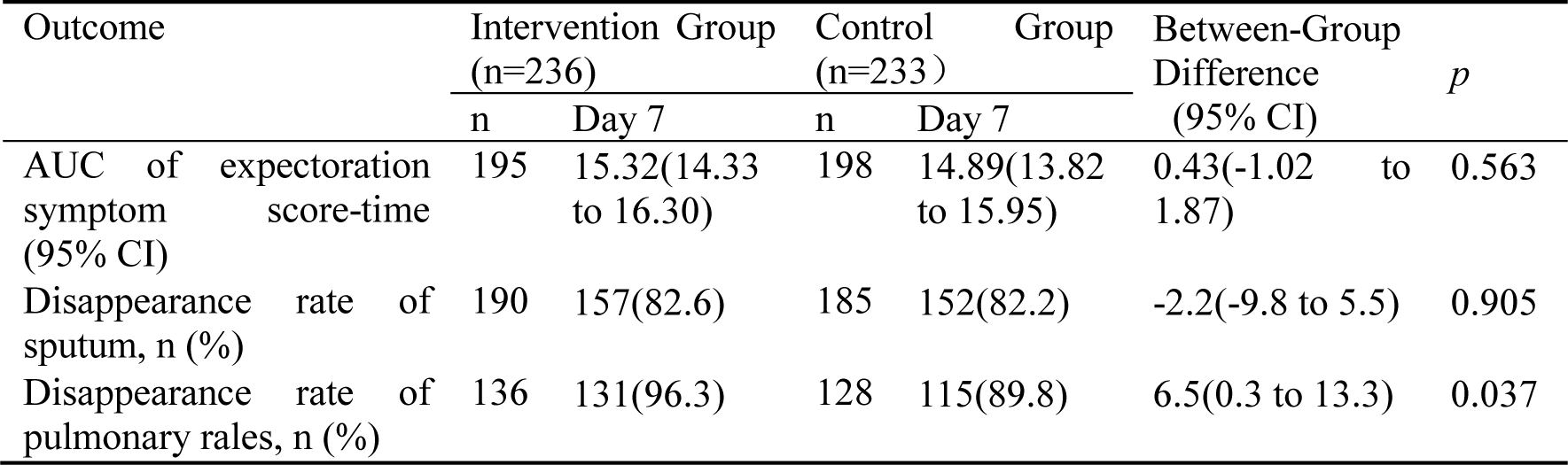
Disappearance rate of secondary clinical symptoms.

**Table 9** lists the results of abnormal laboratory indicators. There were no significant differences in the improvement of abnormally elevated white blood cell count, lymphocyte percentage, and neutrophil percentage between the intervention and control groups. The white blood cell counts in the intervention group exhibited a greater decrease (-2.093 (CI, -3.237 to -0.948)) than the control group (-0.968 [CI, -2.379 to 0.442]) (the difference between groups was -1.12 [CI, -2.87 to 0.62]; *p*=0.199). The lymphocyte percentage in the intervention group exhibited a greater decrease (-8.75 (CI, -12.36 to -5.15)) than the control group (-8.07 [CI, -10.99 to -5.15]) (the difference between groups was -0.68 [CI, -5.29 to -3.93]; *p*=0.770). Besides, the neutrophil percentage in the intervention group exhibited a greater decrease (-21.94 (CI, -29.78 to -14.09)) than the control group (-13.36 [CI, -25.40 to -1.32]) (the difference between groups was -8.58 [CI, -21.49 to -4.33]; *p*=0.181). In comparison to the control group, the intervention group experienced a greater decrease in white blood cell count, lymphocyte percentage, and neutrophil percentage. For abnormally elevated white blood cell count, lymphocyte percentage, and neutrophil percentage, the recovery rates in the intervention group (65.0%, 49.1%, 69.2%) were higher than in the control group (38.5%, 47.3%, 40.0%).

**Table 9.** Abnormal laboratory indicators.

| Outcome | Intervention Group (n=236) | Control Group (n=233) | Between-Group Difference (95% CI) | *p* |
| --- | --- | --- | --- | --- |
| Abnormal increase in white blood cell count |  |  |  |  |
| Cases, n | 20 | 14 | - | - |
| Baseline (SD), 10<sup>9</sup>/L | 10.794(0.777) | 10.731(0.742) | 0.06(-0.48 to 0.61) | 0.813 |
| Day7 (SD), 10<sup>9</sup>/L | 8.702(2.489) | 9.779(2.557) | -1.08(-2.91 to 0.75) | 0.238 |
| Day7-baseline (SD), 10<sup>9</sup>/L | -2.093(2.446) | -0.968(2.334) | -1.12(-2.87 to 0.62) | 0.199 |
| Disappearance, n (%) | 13(65.0) | 5(38.5) | 26.5(-7.3 to 53.3) | 0.135 |
| Abnormal increase in lymphocyte percentage |  |  |  |  |
| Cases, n | 66 | 62 | - | - |
| Baseline (SD), % | 55.27(8.409) | 54.38(6.990) | 0.89(-1.82 to 3.60) | 0.517 |
| Day7 (SD), % | 45.91(11.704) | 46.16(11.417) | -0.25(-4.58 to 4.09) | 0.911 |
| Day7-baseline (SD), % | -8.75(13.593) | -8.07(10.798) | -0.68(-5.29 to 3.93) | 0.770 |
| Disappearance, n (%) | 28(49.1) | 26(47.3) | 1.9(-16.1 to 19.7) | 0.845 |
| Abnormal increase in neutrophil percentage |  |  |  |  |
| Cases, n | 15 | 14 | - | - |
| Baseline (SD), % | 69.50(8.652) | 64.72(7.166) | 4.78(-1.30 to 10.86) | 0.118 |
| Day7 (SD), % | 48.78(8.288) | 49.64(15.089) | -0.86(-11.09 to 9.38) | 0.864 |
| Day7-baseline (SD), % | -21.94(12.980) | -13.36(16.834) | -8.58(-21.49 to 4.33) | 0.181 |
| Disappearance, n (%) | 9(69.2) | 4(40.0) | 29.2(-10.1 to 58.6) | 0.222 |

#### 3.1.4 Safety

The incidence rate of adverse effects was comparable between the intervention group (n=13, 5.5%) and the control group (n=16, 6.9%) (*p*=0.541). The most frequent adverse events (AEs) in both groups were infections and infectious diseases, including urinary tract infection, rhinitis, pulpitis and so on. There were two reported cases of adverse drug reactions (ADRs) in the intervention group, including abdominal pain and vomiting. The severity of these reactions was considered average, and the patients recovered without specific treatment. Neither group of patients reported any severe AEs. Detailed AE records were listed in Supplementary materials (**Table S1, S2**).

### 3.2 Network target analysis of JZOL against AB

To gain insight into the molecular mechanisms of JZOL against bronchitis, analysis on the basis of network target theory and technology^40,41^ was performed. An algorithm based on the drug-gene space was employed to predict the biological effect profiles of the compounds in JZOL. Building upon previous research, the holistic target profile of JZOL was constructed to predict the potential targets of JZOL. It was found that the targets of JZOL mainly fell in 17 pathways or biological processes related to acute bronchitis that could be classified into five modules, in which regulation of immune and inflammation played a leading role in the other four modules, including oxygen metabolism dysfunction, protein metabolism and synthesis, metabolic disorder and cell migration (**Figure 5A**). As the dominant module, the role of immune and inflammation regulation have been reported closely related to AB. T-helper (Th)17 cells and related serum cytokines like IL-17, IL-22 lost their balance in AB according to clinical trials^42^, and some studies have indicated that cytokine responses such as interferon (IFN)-γ, and tumor necrosis factor (TNF)-α decrease with severe RSV disease in infants^43,44^. And in other modules, pathways or biological processes also had been proved to be associated with AB, like oxidative stress^45^, phagocytosis^46^, muscle contraction^47^, leukocyte chemotaxis^48^, protein secretion^49^ and hypoxia^50^. In each module, a biological network was constructed to show the potential network regulation of JZOL on each module during AB, and 42 biomolecules were found to be of vital importance in multi modules. These modular networks might provide important insights for our subsequent molecular mechanism studies of JZOL against AB.

**Figure 5.**
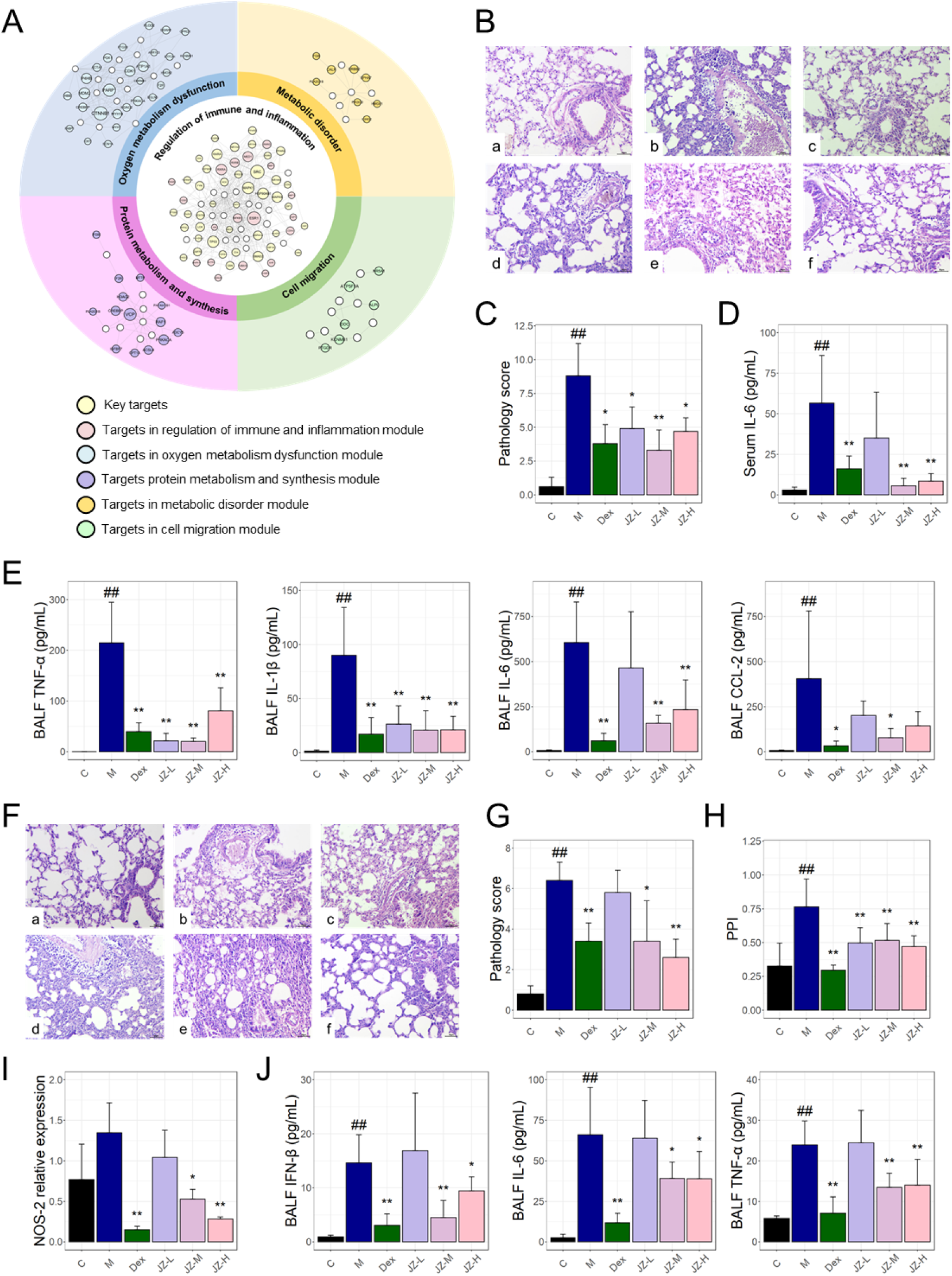
Network target of JZOL in the treatment of cough. (A) Targets network of JZOL against AB. (B) Pathological images of LPS-induced tracheal inflammation animal model. (C) Pathology score of lung tissues in LPS-induced tracheal inflammation animal model. (D) IL-6 level in serum in LPS-induced tracheal inflammation animal model. (E) Levels of TNF-a, IL-1β, IL-6, and CCL-2 in BALF in LPS-induced tracheal inflammation animal model. (F) Pathological images of Poly (I:C)-induced tracheal inflammation animal model. (G) Pathology score of lung tissues in Poly (I:C)-induced tracheal inflammation animal model. (H) PPI in Poly (I:C)-induced tracheal inflammation animal model. (I) NOS-2 level in lung tissues in Poly (I:C)-induced tracheal inflammation animal model. (J) Levels of TNF-a, IL-6, and IFN-β in BALF in Poly (I:C)-induced tracheal inflammation animal model.

Two animal models of inflammation in the trachea (see methods and materials) were constructed to validate the anti-inflammatory effect of JZOL. Both in Lipopolysaccharide (LPS) and Polyinosinic acid-polycytidylic acid (Poly (I:C)) induced animal models of inflammation in the trachea, the infiltration degree of perivascular and alveolar wall inflammation cells in the lungs can be reduced after the intervention of JZOL at middle or high dose (**Figure 5B**, **F**) while the effect of high dose is better. Pathology score of lung tissue was also measured (**Figure 5C**, **G**) and found to decrease significantly after the intervention of JZOL (*p* < 0.05), as well as lung permeability index (PPI) measured in Poly (I:C) induced model (**Figure 5H**). In order to verify the anti-inflammatory effect at molecular level, in LPS induced model, inflammatory cytokines like TNF-α, IL-1β, and CCL-2 were found to be down-regulated significantly by JZOL intervention (**Figure 5E**) in bronchoalveolar lavage fluid (BALF) and IL-6 was decreased after JZOL intervention in both serum and BALF (**Figure 5D**). And in Poly (I:C) induced model, the administration of JZOL at doses of 10 and 20 mL/kg was found to alleviate the infiltration of inflammatory cells in the lung vasculature and alveolar walls, with the higher dose demonstrating a better effect (**Figure 5F**). It was found that the pathology score of lung tissue was also significantly decreased after the intervention of JZOL (**Figure 5G**), consistently with PPI (**Figure 5H**). In molecular level, relative expression level of NOS-2 in lung tissue showed a significant decrease after the treatment of JZOL (**Figure 5I**) while TNF-a, IL-6 and IFN-β in BALF also significantly decreased (**Figure 5J**).

### 3.3 The role of JZOL in the treatment of cough

Further, as core symptom of AB, cough was taken into further consideration. In order to find the mechanism of action of JZOL against cough in AB, we focused the pathways targeted by the holistic targets of JZOL and genes of cough based on network target analysis. Based on the collected molecules related to cough from high level disease-gene association database, pathways related to cough were enriched and classified into different modules (**Figure 6A**). And it was found that IL-17 signaling pathways, Th17 differentiation and TRP channels were in the leading role. Then, with the combination of network target analysis of JZOL and these modular pathways, there are two main modules in the network target of JZOL against cough, including signal transduction and immune system (**Figure 6B**).

**Figure 6.**
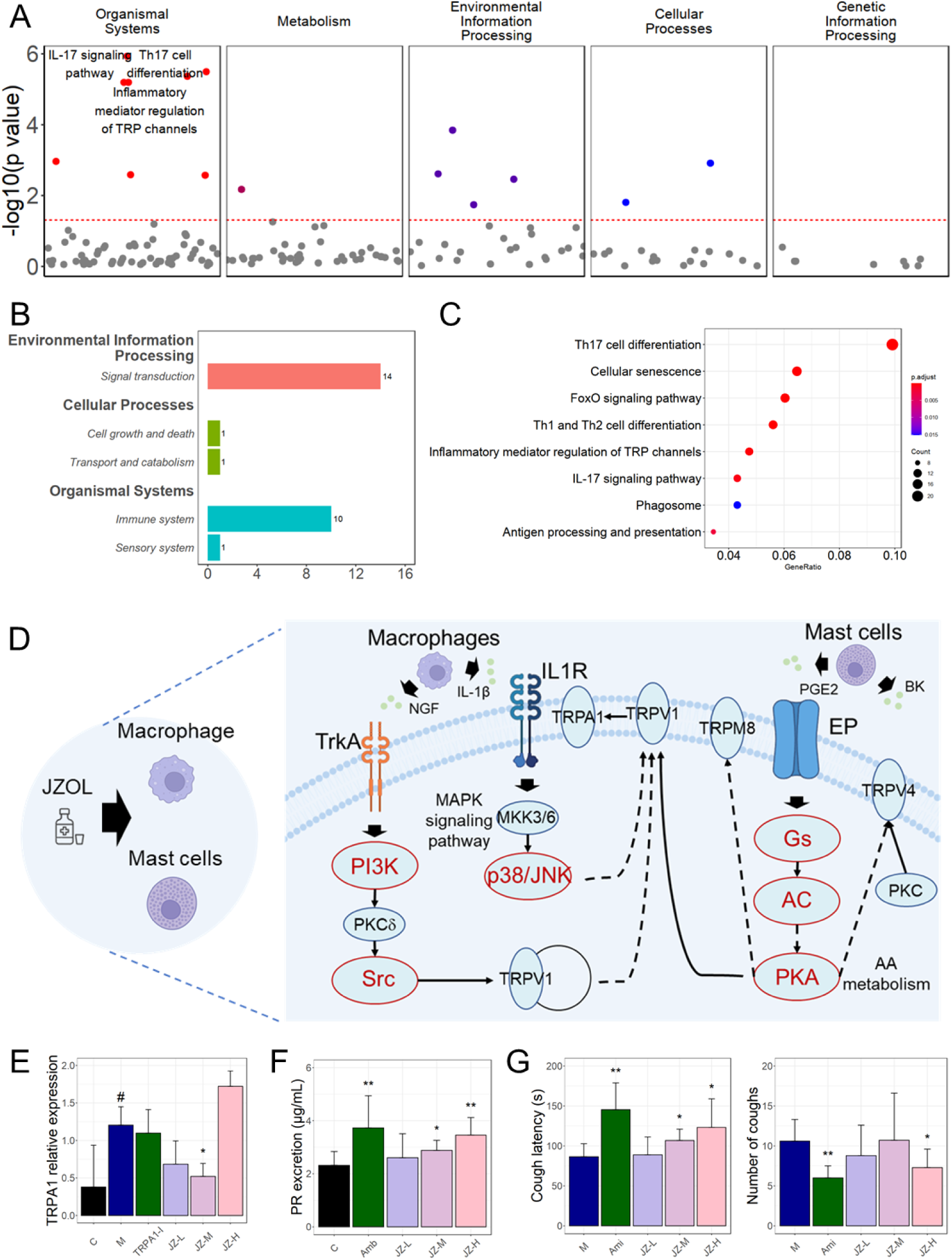
Network target of JZOL in the treatment of cough. (A) Dot plot showing pathways in different classifications enriched by molecules related to cough. (B) Bar plot for predicted genes of JZOL in different modules and corresponding sub-modules. (C) Dot plot showing the potential pathways of JZOL in the treatment of cough. (D) TCM-cell-pathway-molecule multilayer network showing the potential mechanism of JZOL on TRP channels of cough, in which red nodes represent the predicted targets of JZOL in TRP channels. (E) Relative expression level of TRPA1 in tracheal tissues of different animal groups. (F) Phenol red excretion of different animal groups. (G) Cough latency and number of coughs of different animal groups. C: control group, M: model group, TRPA1-I: TRPA1 inhibitor group, Amb: Ambroterol oral solution group, Ami: Aminophylline group, JZ-L: JZOL low dose group, JZ-M: JZOL middle dose group, JZ-H: JZOL high does group.

According to network target of JZOL, inflammatory mediator regulation of TRP channels might play a leading role in the regulation of cough (**Figure 6C**). In this pathway, endogenous inflammatory mediators, including prostaglandin E2 (PGE2) and bradykinin (BK), frequently elevated in respiratory illnesses, were recognized for their ability to induce cough by activating sensory nerves in the airways^51,52^. And TRPA1 and TRP channels were singled out as pivotal regulators of cough responses to both internal and external agents, positioning them as the most promising targets in current efforts to develop anti-cough medications^53^. Based on the result of network target analysis, we have observed that JZOL potentially exerted its regulatory effects on TRPA1 both through direct regulation of relevant targets, as well as indirectly by influencing macrophages and mast cells (**Figure 6D**). Three cellular-molecular pathways were involved in the predicted regulation of JZOL against cough. Two of them starting from macrophage, walked along IL-1β, IL1R, MKK3/6, p38/JNK to TRPV1 and NGF, TrkA, PI3K, PKCδ, Src to TRPV1, respectively. And one from mast cell, started from PGE2 to EP, Gs, AC, cAMP, PKA to TRPV1. Then TRPV1 played a regulatory role on TRPA1. Numerous molecules within the network targeted by JZOL for cough treatment have been previously linked to cough, for example, JNK was a common molecule in many respiratory diseases, like asthma and acute lung injury, and participated in many pathogenic molecular pathways^54–56^. PKA and PKC activation of TRPV1 induced cough and served as an inflammatory marker in evaluating the airway in a postoperative chronic cough model^57,58^. Some of key molecules in these signaling pathways had already been validated in vivo. For example, NGF/TrkA/TRPV1 has been found to be regulated after the intervention of JZOL^21^. These three signaling pathways might potentially offered directions for the validation of molecular-level interactions both in vitro and in vivo during uncovering the mechanism of JZOL against cough for further researches.

In this research, we also conducted three animal models for validating our findings in the treatment of cough with JZOL. Firstly, in cough hypersensitivity animal model which then induced cough with TRPA1 agonist (cinnamaldehyde) stimulation, TRPA1 was found to decrease significantly after JZOL intervention (**Figure 6E**) in the tracheal tissue. And the high-dose JZOL group experienced slight alveolar wall thickening, minimal exudate, and mild local swelling, while the trachea showed dilation without thickening, and there was significant inflammatory cell clustering in the parenchyma. In contrast, the moderate and low-dose JZOL groups had only mild lesions with no significant changes in alveolar walls or exudate (**Figure S1A-F**). Besides, the expectorant effect of JZOL was verified in phenol red excretion model due to the significant increase phenol red excretion after JZOL intervention (**Figure 6F**). As for macroscopic efficacy level, cough latency was significantly increased and number of coughs was significantly decreased (**Figure 6G**) in the treatment with JZOL against citrate acid induced cough model.

Last but not least, in order to observe the regulatory effects of JZOL on TRP channels from a deeper perspective, we have collected transcriptomes for both the representative herbs and compounds in JZOL from ITCM database and previous studies^38,39^. These datasets included the transcriptomes of common compounds in TCM on MCF-7 cell lines and the transcriptomes of common herbs on THP-1 cell lines, respectively, providing transcriptional-level evidence for the influences of representative compounds or herbs of JZOL on the biomolecules of TRP channels to some extent. Firstly, the regulation of representative compounds in herbs of JZOL on biomolecules of TRP channels was measured and representative compounds were determined based on the content determination components recorded in the Chinese Pharmacopoeia. And these compounds have exhibited synergistic intervention effects on the biomolecular of TRP channels (**Figure S2A**). Besides, at the herb’s level, 5 TCM were found in datasets, including Huang Qin, Da Huang, Niu Huang, Shi Gao and Gan Cao. It was found that biomolecules like SRC, GNAS, IL1B, TRPV4, ILR1 and NGF were subject to the synergistic regulatory effects of different herbs in JZOL (**Figure S2B**). And other biomolecules like PRK, JNK (MAPK8 and MAPK10), PI3K were significantly regulated by certain herbs in JZOL.

## 4. Discussion

Acute cough and yellow phlegm are the typical cardinal symptoms of AB. It has been established that the effective ingredients of AHCHOS are ambroxol hydrochloride and clenbuterol hydrochloride, which are representative drugs for symptomatic treatment. AHCHOS, as a mucus motility enhancer, can promote airway cilia oscillation, improve mucus clearance kinetics, and help expel phlegm^55^. According to the Chinese Children’s Medication Guidelines, AHCHOS is recommended for children with wet coughs, including bronchial asthma complicated with infection, persistent bacterial bronchitis, and pertussis^22,60^, and is one of the most important cough and phlegm relieving compound formulations suitable for children. AHCHOS is one of the most representative cough and phlegm reducing western medicines, therefore, it was chosen as the positive control drug for our clinical trial.

Cough is the most common symptom of AB, and the main reason why most children and parents seek treatment is to relieve cough^3,7^. If a cough is not relieved and lasts for more than 3 weeks, it may become chronic. It is now understood that children are more likely to develop recurrent wheezing or asthma than adult bronchitis. More than 80% of children treated in specialized hospitals seek medical attention at least 5 times yearly because of chronic cough^61,62^. Our study indicated that JZOL was not inferior to AHCHOS in treating cough. The median time-to-cough resolution in both groups was 5.0 days, and there was no significant difference among different age groups. After 7 days of treatment, the cough scores of both groups were significantly lower than baseline, and the antitussive onset median time was only 1 day (CI, 1.0d to 2.0d). The cough score, disappearance rate of cough, AUC of cough score-time, and clinical recovery time were similar between the two groups after treatment, suggesting that JZOL can effectively alleviate acute cough in children with AB, and the antitussive effect may not be inferior to β2 receptor agonists, which was contained in AHCHOS. In our cough hypersensitivity and citrate acid induced cough mouse models, we have also demonstrated that JZOL has excellent cough relieving effects and can reduce cough sensitivity.

It is worth noting that the AUC of expectoration symptom score-time and disappearance rate of sputum also showed significant improvement compared to baseline in the two groups, indicating that JZOL has certain advantages in resolving phlegm. Due to the weak cough reflex in children, sputum tends to stay in the airway and block the airway, leading to hypoxemia, hypercapnia, and even respiratory failure. Sputum retention may also aggravate pathological lung changes or secondary new infections, forming a vicious circle with inflammation, obstruction, and further aggravating cough^63,64^. The improvement of expectoration is particularly important for the treatment and prognosis of AB. Our mouse phenol red excretion experiment showed that JZOL can significantly increase the amount of phenol red in bronchial lavage fluid, promote phenol red excretion, further proving the resolving phlegm effect of JZOL.

In clinical study, JZOL and AHCHOS had therapeutic effects on secondary clinical symptoms of AB patients besides cough and expectoration, including fever, thirst, red-faced complexion, dysphoria, short voidings of reddish urine, and dry stool. Although there was no significant difference between the two groups, JZOL yielded a slightly better improvement in several clinical symptoms compared to AHCHOS, including dysphoria and short voidings of reddish urine. Based on the clinical characteristics, the rational use of TCM demonstrated significant clinical value in improving respiratory symptoms caused by AB. The study also substantiated that JZOL could comprehensively treat respiratory and systemic discomfort symptoms caused by AB.

Interestingly, JZOL exhibited the ability to lower abnormally elevated levels of white blood cells, lymphocyte percentage, and neutrophil percentage. Although there were no significant differences between the two groups in the recovery rates of abnormal laboratory indicators, JZOL (65.0%, 49.1%, 69.2%) was significantly higher than those in AHCHOS (38.5%, 47.3%, 40.0%), especially white blood cell count and neutrophil percentage, which indicated that JZOL has potential anti-inflammatory effects. In addition, the disappearance rate of pulmonary rales in the intervention group (96.3%) was significantly higher than the control group (89.8%). In these children, the reduction in pulmonary rales indicated a significant improvement in lung tissue lesions such as bronchial stenosis, airway mucus, or exudate. Given that most children who participated in the trial were too young to explain their symptoms, laboratory indicators, and physical examination were very important for assessing disease recovery. We constructed two animal experiments on tracheal inflammation and evaluated the pathological changes in lung tissue after JZOL intervention. The experimental results indicated that multiple inflammatory factors, such as TNF-α, IL-1β, and IL-6, were downregulated and lung tissue lesions were alleviated after JZOL intervention. The results of our clinical and animal experimental studies all indicated that JZOL may be effective in reducing inflammation and ameliorating damage to lung tissue.

Although AHCHOS is a commonly used western medicine formulation for pediatric AB, its application has many limitations in clinical practice. For example, clenbuterol contained in AHCHOS is a β2 receptor agonist, unless children present wheezing, β2 receptor agonists should not be used to relieve cough routinely^12^. Several reports have shown that clenbuterol has cardiotoxicity and may cause nausea, vomiting, muscle tremors, hypotension, and electrolyte disorders and cannot be used for a long time^65,66^. Although the content of clenbuterol in AHCHOS is within the prescribed safety range, due to the abominable adverse reactions of clenbuterol, AHCHOS is not suitable for young athletes or patients with cardiac insufficiency, electrolyte disorders or require long-term medication treatment. In our clinical trial, we found that the ADRs associated with JZOL were abdominal pain and vomiting, which might be due to the direct absorption of JZOL through the gastrointestinal tract, thus having an impact on the digestive system. Patients recovered spontaneously after discontinuing the medication, and there were no SAEs during the trial. Overall, the incidence of mild to moderate AEs with JZOL (5.5%) was lower than with AHCHOS (6.9%), indicating that JZOL containing “natural” ingredients was safe during medication. JZOL may serve as an alternative for children who cannot use AHCHOS.

Network target analysis^67^ has provided an entry point for studying the mechanism of action of JZOL in the context of AB. It has bridged the gap between clinical phenotypes and microscopic molecular mechanisms. Through network target analysis, we have predicted the modular effects of JZOL in intervening in bronchitis, including its regulatory impact on five modules, in which regulation of immune and inflammation played a leading role in the other four modules, including oxygen metabolism dysfunction, protein metabolism and synthesis, metabolic disorder and cell migration. Combined with clinical trials, the mechanism of JZOL against cough in AB was uncovered by network target analysis. At the core pathway--inflammatory mediator regulation of TRP channels, we delineated the mechanism of JZOL’s intervention on the cough-related target TRPA1 at molecular pathway level. And three pathways were found to be of vital importance during the intervention of JZOL, including IL-1β/IL1R/TRPV1/TRPA1 and NGF/TrkA/TRPV1/TRPA1 started from macrophages and PGE2/EP/PKA/TRPV1/TRPA1 from mast cells. We also conducted in vivo experiments to verify our findings. With these findings and corresponding validations, JZOL was proved to be able to improve the pathology score of lung tissue, reduce lung permeability, down-regulate the expression of various inflammatory factors such as TNF-α and IL-1β, while also prolonging the latent period of cough, reducing the number of coughs, and regulating the expression levels of some key molecules in the TRP channels.

Our study still has limitations. First of all, in clinical trial, we did not conduct long-term follow-ups. Children under 2 years old were not included in the study. As infants under the age of 2 years have an underdeveloped respiratory system and for the children’s safety and the psychological acceptance of their guardians, we did not observe the treatment effect in this age group. Considering the rapid progression and low self-healing rate of AB, as well as the prognostic risk associated with children, our clinical trial design did not include a control group of children who received no intervention or placebo. Instead, we compared the treatment outcomes of children using the positive drug AHCHOS. Indeed, the study’s main purpose was to evaluate the efficacy of JZOL in treating cough and phlegm with AB. In further research, to investigate the molecular mechanisms of JZOL in treating acute childhood bronchitis, we plan to conduct experimental validations composed of clinical multi-omics and drug-target relationships. And these findings in this study will provide a targeted focus of our future research endeavors.

In summary, we found that the efficacy of JZOL in treating AB is not inferior to AHCHOS, and it is a safe and effective intervention to improve the clinical symptoms of AB. Based on clinical findings, we conducted network target analysis and animal experiments, further verified that JZOL had a vital importance role on inflammation and immune regulatory. And pathways in TRP channels like IL-1β/IL1R/TRPV1/TRPA1, NGF/TrkA/TRPV1/TRPA1 and PGE2/EP/PKA/TRPV1/TRPA1 were important in the treatment with JZOL against cough.

## Acronyms

AB: acute bronchitis
CAM: Complementary and Alternative Medicine
TCM: Traditional Chinese medicine
JZOL: Jinzhen Oral Liquid
AHCHOS: Ambroxol Hydrochloride and Clenbuterol Hydrochloride Oral Solution
AUC: Area Under Curve
HR: hazard ratio
ITT: intention-to-treat
FAS: full analysis set
KEGG: Kyoto Encyclopedia of Genes and Genomes
GO: Gene Ontology
AEs: adverse events
ADRs: adverse drug reactions
Th: T-helper
IFN: interferon
TNF: tumor necrosis factor
LPS: lipopolysaccharide
Poly (I:C): polyinosinic acid-polycytidylic acid
PPI: permeability index
BALF: bronchoalveolar lavage fluid
PGE2: prostaglandin E2
BK: bradykinin.

## Contributors

SXW and SL designed the study, did the data interpretation, and revised the manuscript. QHF, YWD, BYW, CMW, YMX, ZLZ, WQS, ZZW, CCX and XKL for assistance in careful editing of the manuscript and JHW, YD, XJA, JC, YYX, RY, NL, JW, YQT, HFL, NY, YLW, XLH, WP, YFL, XQX, QYZ, CSL, JX, HTZ, LZ, QQR, YX and QS, who participated in collection of clinical data. BYW performed network target analysis. All coauthors approved the final version of the manuscript.

## Data sharing statement

The data collected in this study are available to researchers through QinHua Fan. Data requests will be reviewed and approved on the basis of scientific merit, and with a signed data access and sharing agreement. These data will be available for a minimum of 5 years after publication.

## Acknowledgements

The authors acknowledge Jiangsu Kangyuan Pharmaceutical Co., Ltd for providing medication and placebo, thank the patients for participating in this study, and the research staff at each hospital site for data collection, thank Shanghai Bojia Pharmaceutical Technology Co., Ltd for statistical analysis, and also thank Home for Researchers editorial team (www.home-for-researchers.com) for language editing service.

