## Supplementary materials for "Depicting the regulatory role of JZOL on TRP channels in the treatment of Acute Bronchitis based on the combination of clinical trials, computational analysis and in vivo experiments"

### Methods and Materials

### LPS-induced tracheal inflammation animal models

C57BL/6J mice, male, weighing 18-20g, were purchased from Sibefo (Beijing) Biotechnology Co., Ltd. with a qualification certificate number of No.1103242011013807 and a production license number of SCXK (Jing) 2019-0010. The rearing environment was a barrier system with a license number of SYXK (Su) 20180026, room temperature at 20-22℃, relative humidity between 40-70%, and a light cycle of 12h/12h. The experimental animals were maintained with feed purchased from Jiangsu Xietong Pharmaceutical Bioengineering Co., Ltd., with a production license number of Su Feeding Certificate (2019) 01003.

Sixty male C57BL/6J mice weighing 18-20g were divided into the following groups: normal group, model group, dexamethasone group (0.195mg/kg), and Jinchun Oral Liquid groups at low, medium, and high doses (5, 10, 20 mL/kg). Isoflurane gas anesthesia was used, and 40μL of LPS (1mg/mL) was instilled into the nostrils of the mice, maintaining an upright position for 1 minute. The mice were modeled for 3 consecutive days. Concurrently with modeling, each group was administered the corresponding drug by gavage; the model control group was given the same volume of distilled water (10 mL/kg body weight) by gavage once a day for 5 consecutive days. One hour after the last administration, blood was collected from the eyes of the animals, centrifuged to obtain serum, and a tracheal cannula was inserted via a neck surgery to lavage the lungs 3 times with 0.5mL of PBS, collecting the bronchoalveolar lavage fluid (BALF). The right lower lung was sampled, fixed with 4% formaldehyde, stained with HE to observe histopathological changes. Additional lung tissue was collected for Western Blot analysis. All samples to be tested were stored at -80°C for freezing.

### Poly (I:C)-induced tracheal inflammation animal models

Sixty male C57BL/6J mice weighing 18-20g were obtained from Sibefo (Beijing) Biotechnology Co., Ltd., with a qualification certificate number of No.1103242011007298 and a production license number of SCXK (Jing) 2019-0010. The mice were housed in a barrier system with a license number of SYXK (Su) 20180026, at a room temperature of 20-22℃, relative humidity between 40-70%, and a 12-hour light/dark cycle. The experimental animals were maintained with feed purchased from Jiangsu Xietong Pharmaceutical Bioengineering Co., Ltd., with a production license number of Su Feeding Certificate (2019) 01003.

The mice were divided into the following groups: normal group, model group, dexamethasone group (0.195mg/kg), and Jinchun Oral Liquid groups at low, medium, and high doses (5, 10, 20 mL/kg). All animals were administered the corresponding drugs via gavage. The normal group and model group were given physiological saline, with an administration volume of 20 mL/kg, once a day for 7 consecutive days. On days 4-6, the mice in each group were anesthetized with isoflurane and laid supine, then 40μL of Poly(I:C) saline solution (1mg/mL) was slowly instilled into the nostrils of the mice; the normal group was given an equal volume of saline, once a day for 3 consecutive days. On day 7, one hour after the last administration, blood was collected from the eyes of the mice, and serum was obtained by centrifugation. The neck was then incised, the trachea was exposed, and a catheter was inserted through a small incision at the upper end of the trachea and secured. The chest was opened, the right lung lobe was ligated, and the left lung was lavaged with 0.9mL of PBS three times, each time with 0.3mL, aspirated three times, with a total recovery rate of 60%~70%. The bronchoalveolar lavage fluid (BALF) was centrifuged at 5000rpm for 10 minutes, and the supernatant was collected for the detection of inflammatory factors and lung alveolar permeability index PPI (measuring the protein concentration in BALF and serum, calculating the ratio). The upper lobe of the right lung was taken for pathological examination; the middle lobe of the right lung was fixed in 4% formaldehyde for pathological examination; and the lower lobe of the right lung was taken for Western Blot, qPCR, and other tests. All samples to be tested were stored at -80°C for freezing.

### Phenol red excretion animal models

ICR mice, SPF grade, with an equal number of males and females, weighing 18-20g, were provided by the Experimental Animal Center of Nantong University, with an animal production license number: SYXK(Su) 2019-0001, and an animal qualification certificate number: NO.202002007. The mice were housed in a barrier system with a license number: SYXK(Su)20180026, at a room temperature of 20-22℃, relative humidity of 40-70%, and a 12-hour light/dark cycle. The experimental animals were maintained with feed purchased from Jiangsu Xietong Pharmaceutical Bioengineering Co., Ltd., with a production license number: Su Feeding Certificate (2019) 01003.

Fifty mice, with an equal number of males and females, were randomly divided into a blank group, ambroxol oral solution group (10.4 mL/kg), and Jinchun Oral Liquid groups at low, medium, and high doses (5, 10, 20 mL/kg). All animals were administered the corresponding drugs via gavage; the blank group was given distilled water, once a day for 7 consecutive days. The mice were fasted for 16 hours before the experiment but had access to water. One hour after the last administration, the mice were intraperitoneally injected with 1% phenol red solution at a dose of 50 mL/kg (prepared by dissolving 1.0g of phenol red in 5ml of 1mol/L NaOH, followed by dilution to 100ml with normal saline). Thirty minutes after the injection, the animals were euthanized, positioned supine, and the neck skin was incised to expose the trachea. A flattened 7-gauge needle was inserted into the trachea approximately 0.3cm and secured with silk thread. A syringe was used to instill 0.5ml of 5% sodium bicarbonate solution into the respiratory tract through the needle, and the lavage was performed three times. The lavage fluid was collected in a test tube, and the process was repeated twice more, with a total combined lavage volume of at least 1.2mL, which was then centrifuged and set aside for testing.

Phenol red was precisely weighed to 50mg, dissolved in 1mol/L NaOH solution, and then sequentially diluted with 5% sodium bicarbonate solution to concentrations of 10.00ug/ml, 5.00ug/ml, 2.50ug/ml, 1.00ug/ml, and 0.50ug/ml. Absorbance was measured at 546nm, and a standard curve was created with concentration on the x-axis and absorbance on the y-axis. The lavage fluids from each group were colorimetrically analyzed at 546nm to determine absorbance values. Based on the phenol red standard curve, the excretion amount and increase rate of phenol red were calculated.

### Citric acid induced cough animal models

Guinea pigs, conventional grade, weighing 260-290g, with an equal number of males and females, were obtained from Lai Fu Breeding Farm in Pukou District, Nanjing City. The animal production license number is SYXK(Su) 2019-0005, and the qualification certificate number is NO.202017045. The animals were housed in polycarbonate plastic boxes, with males and females kept separately, with 10 animals per sex per cage. They were maintained under conventional environmental conditions with an experimental animal use license number: SYXK(Su) 2018-0025, applicable to the conventional environment. Animals had free access to water during the quarantine/domestication period and the experimental period.

The guinea pigs were allowed to adapt to the housing environment for 7 days. Preliminary screening: Healthy guinea pigs weighing 220-240g were placed in an ultrasonic nebulizer and observed for cough latency (the time from the start of the 17.5% citric acid injection to the onset of coughing), which should be less than 2 minutes for qualification.

Qualified guinea pigs were randomly assigned to groups based on the number of coughs: model control group, aminophylline group (46.5mg/kg), and Jinchun Oral Liquid groups at low, medium, and high doses (2.32, 4.64, 9.28 mL/kg), with 10 animals in each group, half males, and half females. The animals were administered the corresponding drugs via gavage; the model control group was given the same volume of distilled water (10 mL/kg body weight) via gavage once a day for 7 consecutive days. One hour after the last administration, the guinea pigs were placed in a 4L sealed glass box, and 17.5% citric acid solution was evenly sprayed into the box using an ultrasonic nebulizer for 30 seconds. The effects of the drugs on cough latency and the number of coughs within 5 minutes were recorded.

### Cough hypersensitivity animal model

Sixty-eight-week-old BALB/c mice (purchased from Sibefo (Beijing) Biotechnology Co., Ltd.) were used, all female, SPF grade, weighing 20g-22g. The animal license number is SCXK (Jing) 2019-0010. The mice were fed with regular feed and kept at a room temperature of 22℃-24℃, with a relative humidity of 58% and a 12-hour light/dark cycle. They had free access to food and water.

After a 3-day adaptation period, 9 mice were randomly selected as the blank control group, and the remaining mice were established with a cough hyper-sensitivity model. The procedure involved: after adaptation to the environment, on the first day, the mice were intraperitoneally injected with 0.1ml of ovalbumin sensitization solution (containing 100ug of ovalbumin and 400ug of aluminum hydroxide gel), then re-sensitized on the 7th and 14th days, and starting from the 21st day, they were stimulated with 1% ovalbumin solution via ultrasonic nebulization for 30 minutes daily, lasting for 3 days.

On the 24th day, the successfully modeled mice were randomly divided into groups and administered medication. The groups included: model group, JZOL high-dose group, JZOL medium-dose group, JZOL low-dose group, and TRPA1 inhibitor group, with 9 mice in each group. The drug doses for each group were calculated based on the recommended daily dose for adults, adjusted for body weight. The TRPA1 inhibitor group received 2 mmol·L-1 HC-030031, which was inhaled daily for 5 minutes. The model group received distilled water by gavage. The JZOL high, medium, and low-dose groups received 24.64mL·kg-1, 12.32mL·kg-1, and 6.16mL·kg-1 JZOL, respectively, which were dissolved in distilled water and administered orally. Each group received treatment once a day for 5 consecutive days.

On the 28th day, each group was given a TRPA1 agonist (cinnamaldehyde) to induce coughing. Cinnamaldehyde 50mg was added to 1 mL of Tween-80 solution and 1 mL of absolute ethanol, followed by the addition of 8.0mL of normal saline to prepare a 10-2mol·L-1 cinnamaldehyde stock solution. 2mL of the cinnamaldehyde stock solution was diluted to 50mL to create a 2×l0-4mol·L-1 solution. The mice were placed in a transparent wide-mouth plastic bottle and exposed to the cinnamaldehyde aqueous solution steam for 15 seconds using an ultrasonic nebulizer to induce coughing.

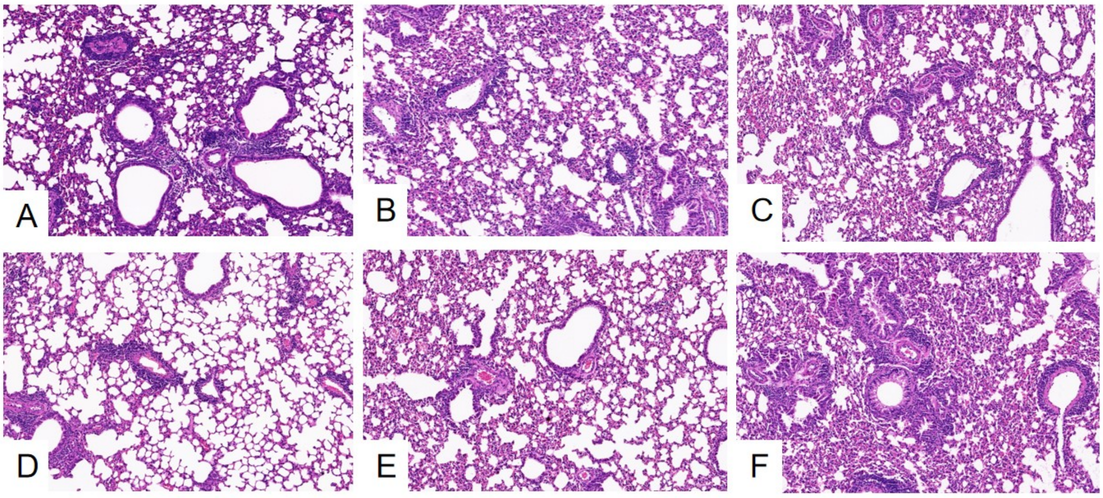


***Figure S1.*** Pathological images of cough hypersensitivity animal model. (A) Model group. (B) Control group. (C) JZOL high dose group. (D) JZOL middle dose group. (E) JZOL low dose group. (F) TRPA1 inhibitor group.

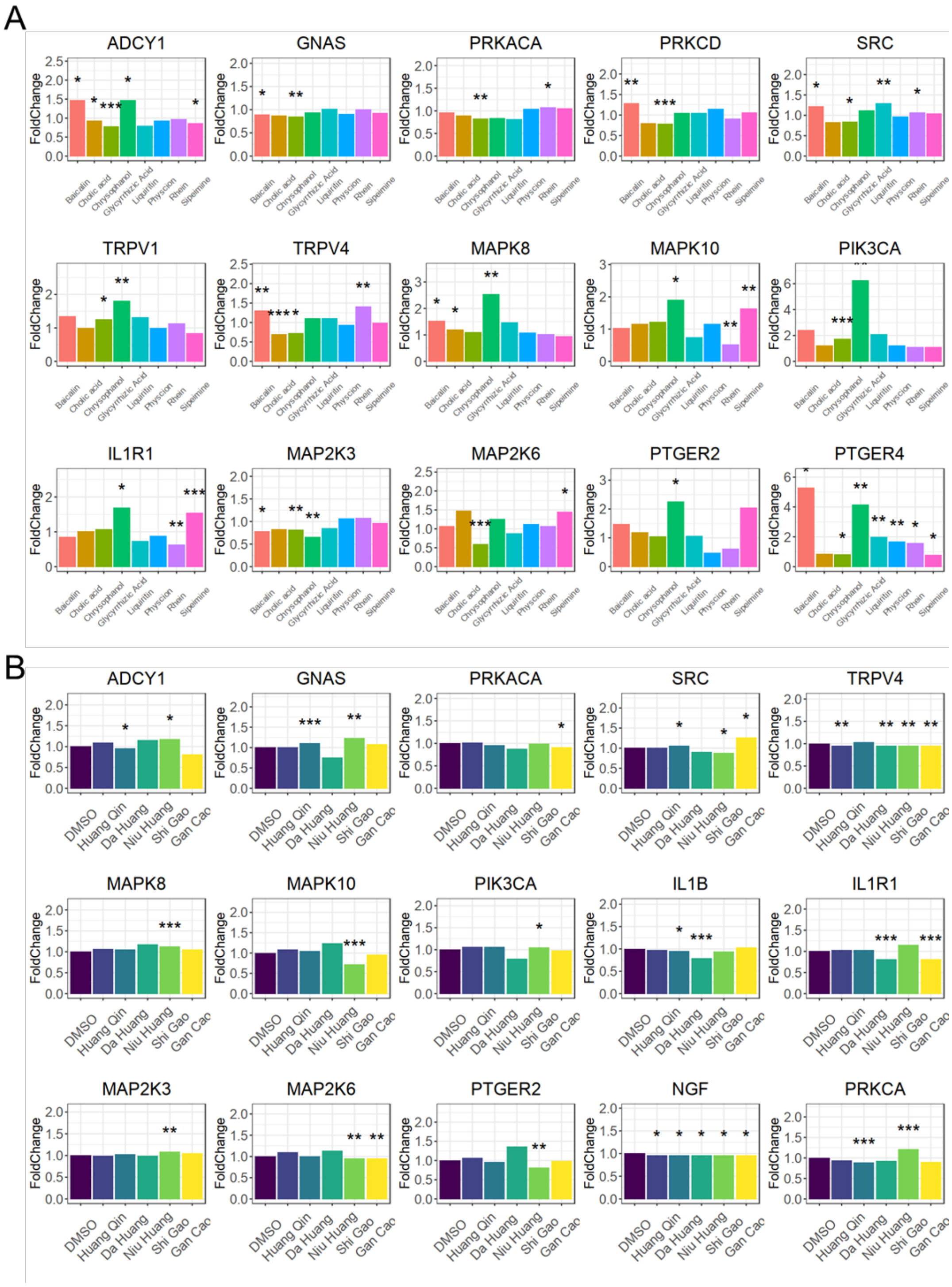


***Figure S2.*** Public transcriptomics validation of the regulation on key biomolecules in TRP channel of representative herbs and compounds in JZOL. (A) Public validation-based bar plot showing the fold change and statistical significance of the regulatory effect on the expression of biomolecules of TRP channels by representative compounds in JZOL. (B) Bar plot based on public validation data, illustrating the fold change and statistical significance of the effects on TRP channels biomolecule expression regulated by representative herbs in JZOL.

***Table S1.*** The Incidence Rate of TEAE (SS)

| | Intervention Group (n=235) | Control Group (n=232) | *p* |
|---|---|---|---|
| TEAE | 13(5.5) | 16(6.9) | 0.541 |
| ADR | 1(0.4) | 0(0) | >0.999 |
| SAE | 0(0) | 0(0) | NA |
| SADR | 0(0) | 0(0) | NA |
| TEAE leading to dose reduction | 0(0) | 0(0) | NA |
| ADR leading to dose reduction | 0(0) | 0(0) | NA |
| TEAE causing interruption of use | 0(0) | 1(0.4) | 0.497 |
| ADR causing interruption of use | 0(0) | 0(0) | NA |
| TEAE causing complete cessation of use | 1(0.4) | 1(0.4) | >0.999 |
| ADR causing complete cessation of use | 0(0) | 0(0) | NA |
| TEAE leading to withdrawal from the trial | 1(0.4) | 1(0.4) | >0.999 |
| ADR leading to withdrawal from the trial | 0(0) | 0(0) | NA |
| TEAE causing death | 0(0) | 0(0) | NA |
| ADR causing death | 0(0) | 0(0) | NA |
| III-V level TEAE | 0(0) | 1(0.4) | 0.497 |
| III-V level ADR | 0(0) | 0(0) | NA |
| Important TEAE | 7(3.0) | 7(3.0) | 0.981 |
| Important ADR | 0(0) | 0(0) | NA |

The relationship with the investigational drug is positively related, likely related, possibly related, pending evaluation, and cannot be evaluated, all of these are considered drug related TEAEs. TEAE:Treatment Emerged Adverse Event, SS: Secure Set, ADR: Adverse Drug Reactions, SAE: Serious Adverse Event, SADR: Serious Adverse Drug Reactions.

*Table S2.* TEAE records (SS)

| SOC | Intervention Group (n=235) | | Control Group (n=232) | |
|---|---|---|---|---|
| PT | Number | Frequency | Number | Frequency |
| **Total** | 13(5.5) | 15 | 16(6.9) | 17 |
| **Infectious diseases** | 4(1.7) | 4 | 6(2.6) | 6 |
| urinary tract infection | 0(0) | 0 | 3(1.3) | 3 |
| upper respiratory tract infection | 2(0.9) | 2 | 0(0) | 0 |
| Nasopharyngitis | 0(0) | 0 | | |
| rhinitis | 0(0) | 0 | 1(0.4) | 1 |
| virus infection | 0(0) | 0 | 1(0.4) | 1 |
| Bacterial infection | 1(0.4) | 1 | 0(0) | 0 |
| pulpitis | 1(0.4) | 1 | 0(0) | 0 |
| **Laboratory Examination** | 3(1.3) | 3 | 3(1.3) | 4 |
| urinary white blood cell positivity | 0(0) | 0 | 1(0.4) | 1 |
| urinary leukocyte esterase positive | 0(0) | 0 | 1(0.4) | 1 |
| urinary sediment detection | 0(0) | 0 | 1(0.4) | 1 |
| urinary protein detection | 1(0.4) | 1 | 0(0) | 0 |
| urinary occult blood positive | 1(0.4) | 1 | 0(0) | 0 |
| electrocardiogram T-wave apex | 1(0.4) | 1 | 0(0) | 0 |
| abnormal electrocardiogram | 0(0) | 0 | 1(0.4) | 1 |
| **Systemic diseases and various reactions at the administration site** | 2(0.9) | 2 | 4(1.7) | 4 |
| fever | 2(0.9) | 2 | 4(1.7) | 4 |
| **Respiratory, thoracic, and mediastinal diseases** | 2(0.9) | 2 | 1(0.4) | 1 |
| allergic rhinitis | 1(0.4) | 1 | 0(0) | 0 |
| cough | 1(0.4) | 1 | 0(0) | 0 |
| Oropharyngeal pain | 0(0) | 0 | 1(0.4) | 1 |
| **Metabolic and nutritional diseases** | 0(0) | 0 | 1(0.4) | 1 |
| anorexia | 0(0) | 0 | 1(0.4) | 1 |
| **Various neurological diseases** | 1(0.4) | 1 | 0(0) | 0 |
| headache | 1(0.4) | 1 | 0(0) | 0 |
| **Various types of injuries, poisoning, and operational complications** | 1(0.4) | 1 | 0(0) | 0 |
| fracture | 1(0.4) | 1 | 0(0) | 0 |
| **Congenital familial hereditary diseases** | 0(0) | 0 | 1(0.4) | 1 |
| phimosis | 0(0) | 0 | 1(0.4) | 1 |
| **Gastrointestinal system diseases** | 1(0.4) | 2 | 0(0) | 0 |

| | | | | |
|---|---|---|---|---|
| abdominal pain | 1(0.4) | 1 | 0(0) | 0 |
| vomit | 1(0.4) | 1 | 0(0) | 0 |

MedDRA 25.0 was used to encode. SOC and PT were arranged in descending order according to the total number of AE cases. Number, one subject was counted up to once in the same term (SOC or PT) at most. Frequency, one subject was counted up the actual number of occurrences in the same term (SOC or PT). SOC: System Organ Classification, PT: Preferredterm.
